# *ASXL3* truncating patient variants mediate transcriptional gain-of-function and are antisense oligonucleotide-responsive

**DOI:** 10.64898/2026.07.20.26358515

**Authors:** Yuji Nakamura, Toan Nguyen, Nofar Mor, Dan Dominissini, Isaac Tang, Christopher Jay Torio, Harikrishna Thulaseedharan, Rachel Yibei Zhou, Wenshu Zhang, Konstantina Skourti-Stathaki, Julie Douville, Laurence Mignon, Ashley Dung, Susan Freier, Andrew Watt, Sujatha Jagannathan, Stanley T. Crooke, Joseph G. Gleeson

## Abstract

**One Sentence Summary:** *ASXL3* patient truncations cause BRS through GOF protein accumulation, not LOF, and ASO-mediated knockdown reverses disease transcriptional signatures.

Truncating variants in the human Additional sex combs (*asx*) *ASXL* genes are frequent in clonal hematopoiesis and severe dominant neurodevelopmental syndromes, yet are assumed to represent loss-of-function (LOF) alleles. However, numerous LOF alleles are documented in healthy individuals. Here we show *ASXL3* patient truncations in neurodevelopmental condition Bainbridge-Ropers syndrome (BRS), by virtue of their distinct location in the gene body, instead mediates gain-of-function (GOF) by escaping nonsense-mediated decay and Cullin 4-dependent degradation, resulting in aberrant protein accumulation, widespread transcriptional dysregulation, and altered chromatin accessibility. Patient-derived cell lines partnered with CRISPR knock-in of patient versus population truncations excluded simple haploinsufficiency, and instead support this ‘two-hit’ GOF mechanism. Deletion mapping identified a broad C-terminal destabilizing region, explaining the 3′ clustering of benign truncations. Haploinsufficiency was further excluded by forced expression of full length *ASXL3* in patient lines, which failed to rescue the disease-associated differential gene expression signature. By contrast, antisense oligonucleotides targeting *ASXL3* largely normalized this signature, providing a mechanistic rationale for knockdown therapies in *ASXL* associated disease.

## Introduction

Additional sex combs-like *ASXL* genes *ASXL1-3* encode chromatin-associated scaffold proteins that integrate Polycomb and Trithorax group activities to regulate the epigenome, impacting transcription, development and tissue homeostasis. As human homologs of Drosophila *Asx*, they assemble with the deubiquitinase BAP1 in the PR-DUB complex to remove H2AK119 monoubiquitin and, in partnership with PRC2, contribute to H3K27 trimethylation at developmental loci(*1, 2*). *ASXL1* and *ASXL2* are broadly expressed, whereas *ASXL3* is enriched in the brain, ovary, and testis(*3*). Germline truncating variants in each paralog cause distinct but overlapping neurodevelopmental conditions, including Bohring–Opitz (OMIM: *ASXL1*), Shashi–Pena (OMIM: *ASXL2*), and Bainbridge–Ropers (OMIM: *ASXL3*) syndromes(*4*). In parallel, recurrent somatic truncating *ASXL1* and *ASXL2* variants act as key drivers in clonal hematopoiesis and myeloid malignancies(*5*), underscoring the broad biological and clinical importance of *ASXL* dosage and domain integrity.

Truncating variants dominate the human pathogenic spectrum of *ASXL* genes. By analogy to biallelic loss of *Asx* in flies and *Asxl1* mouse knockouts, they have long been interpreted as classical LOF alleles(*1, 6*). This view has shaped clinical practice, with most *ASXL* nonsense, frameshift, and splice variants introducing premature termination codons (PTCs) classified as null, despite the numerous early truncations documented as tolerated in population cohorts predicted to undergo nonsense-mediated decay (NMD)(*7*). The resulting paradoxical discrepancy between the high burden of *ASXL* truncations in ostensibly healthy individuals, with severe phenotypes seen in patients, has fueled debate over penetrance, created substantial uncertainty for variant interpretation, prompted ad hoc hypotheses about genetic modifiers, and muddied genetic counselling and therapeutic approaches(*8*).

Among the many coding genes in the human genome, recent evidence suggests a diversity of outcomes resulting from truncating variants, in part mediated by NMD evasion, with additional factors regulating function. For instance, *NARS1* Asparaginyl-tRNA synthetase-1 shows late truncating dominant variants that evade NMD as well as early truncating recessive variants, both associated with nearly identical pediatric neurodegeneration(*9*). The likely explanation is that dominant mutations evade NMD to yield a toxic protein that interferes with function of the healthy copy. Other genes such as *POGZ* and *MN1* show phenotypes that require the transcripts to evade NMD(*10, 11*).

While these examples follow standard NMD evasion rules (i.e. last exon, 50-55nt penultimate exon)(*12*), recent saturation genome editing demonstrates that NMD activity may vary quantitatively with PTC position and local sequence context, rather than following simple binary rules(*13*). Thus, the PTC position within the gene body can critically shape transcript stability and protein abundance. Here, we show that late but not early *ASXL3* truncations similarly escape NMD to create stable proteins. These truncations lack a critical C-terminal degron signal that leads to toxic effects. This ‘two-hit’ model has potential therapeutic implications.

## Results

### PTC position correlates with pathogenicity for *ASXL3* truncations

To understand the *ASXL3* truncation paradox, we evaluated locations of 220 truncations in ClinVar and 90 truncations in gnomAD, representing diseased- and healthy-individuals, respectively. *ASXL3* contains 10 upstream relatively short exons, followed by two large terminal exons of 1,957 and 3,708 coding bases, respectively. Notably, ClinVar truncations were enriched in the 5′ region of the last two exons. In contrast, gnomAD truncations were distributed across the 10 upstream exons and the distal 3′ end of exon 12 (residues ∼1,900–2,248, Fig. 1A)(*4*). To investigate the susceptibility of *ASXL3* truncating variants to NMD, we applied the machine learning-based NMD prediction algorithm NMDetectiveAI, which incorporates quantitative NMD rules such as gradients across long exons, as described for *LMNA*(*13*). In *ASXL3*, in contrast to the uniform NMD susceptibility predicted by the classical binary rule, NMDetectiveAI predicted an NMD gradient across the penultimate exon, mirroring the progressive decrease in ClinVar variants toward the 3′ end (Fig. 1A).

**Fig. 1.**
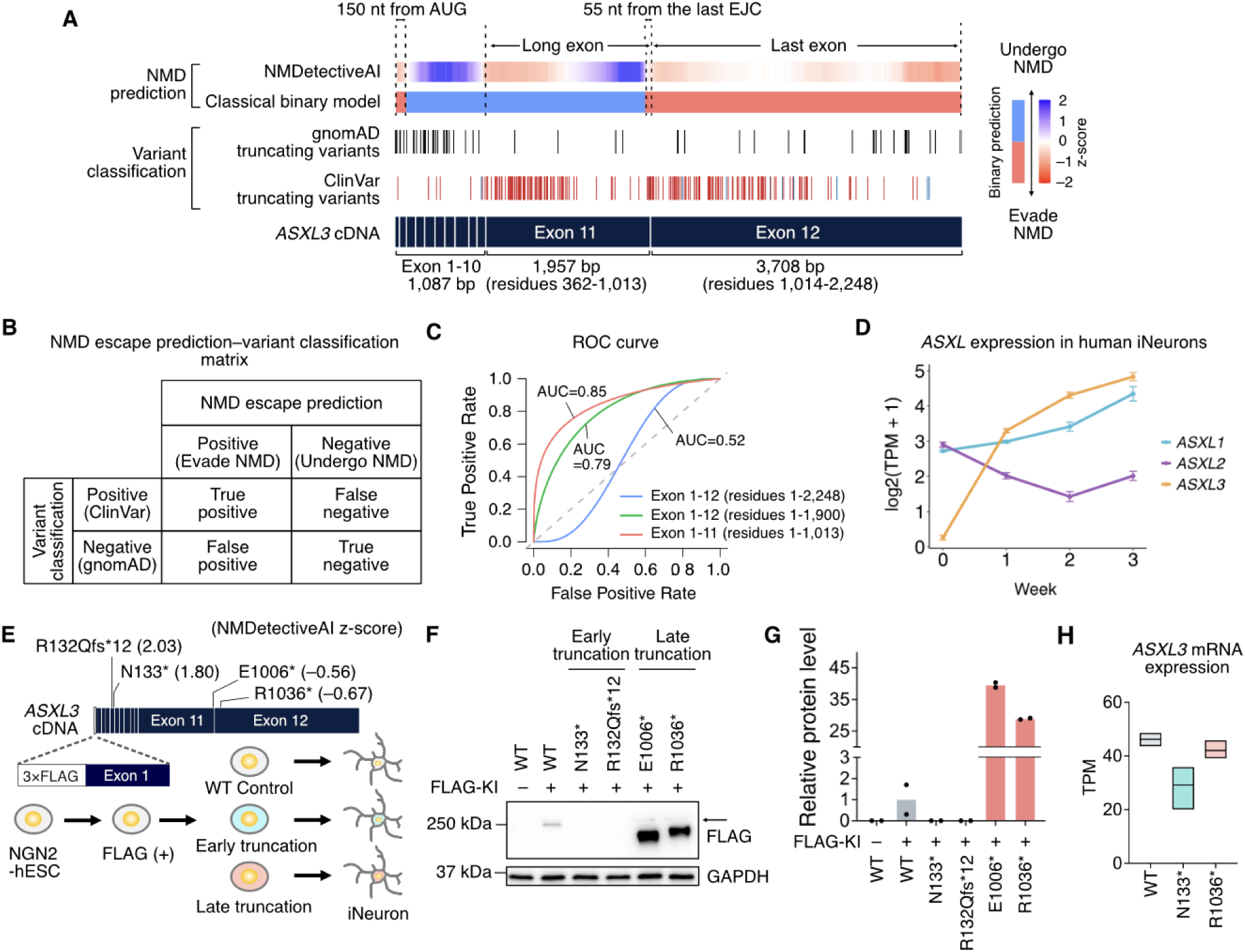
Patient but not control *ASXL3* truncations evade NMD and produce aberrantly accumulated ASXL3 protein in iNeurons. (**A**) NMD prediction at various PTCs in *ASXL3*. PTCs from gnomAD (black) and ClinVar (pathogenic or likely pathogenic, red; variants of uncertain significance, blue). NMD behavior across the coding sequence was predicted using NMDetectiveAI and a classical binary model. For NMDetectiveAI, prediction scores shown as z-scores. Higher values: greater likelihood of undergoing NMD. (**B**) Confusion matrix of ClinVar versus gnomAD truncating variants based on predicted NMD escape using NMDetectiveAI. Variants from ClinVar predicted to escape NMD defined as TP, whereas variants from gnomAD predicted to undergo NMD defined as TN. Variants falsely predicted defined as FN and FP. (**C**) ROC curves with AUC values for NMD-based classification of ClinVar and gnomAD variants. Performance was near random when all PTCs were included (blue), but improved upon excluding variants in exon 12 or beyond residue 1,900 (red and green). These data suggest that NMDetectiveAI accurately predicts NMD status except for exon 12, and that there may be an additional factor in exon 12 contributing to pathogenicity. (**D**) Temporal expression of *ASXL* paralogues during WT iNeurons differentiation (*n* = 4 biological replicates). Error bars: SEM. (**E**) CRISPR-engineering of H9 NGN2-hESCs. A 3×FLAG tag was knocked in to 5’ region of *ASXL3*. Early and late *ASXL3* truncations were introduced into H9 NGN2-hESC lines to assess endogenous *ASXL3* expression. NMDetectiveAI z-scores shown for the indicated truncations. (**F**) Western blot of DIV15 iNeurons analyzed for FLAG-tagged ASXL3 demonstrating PTC position-dependent protein abundance. GAPDH: loading control. Arrow: nonspecific band. (**G**) Quantification of FLAG-tagged ASXL3 (normalized to GAPDH). Fold change relative to FLAG KI WT, with >25-fold increases in E1006* and R1036*. Biological replicate (*n* = 2). Break in y-axis was used to accommodate all variants. (**H**) RNA-seq of DIV15 iNeurons assessing *ASXL3* mRNA expression, consistent with NMD of a representative early truncation and NMD escape of a representative late truncation. *n* = 3 biological replicates. Floating bars (min to max) with the mean indicated by a horizontal line. Abbreviations: AUC, area under the curve; DIV, days in vitro; EJC, exon junction complex; KI, knock-in; PTC, premature termination codons; ROC, receiver operating characteristic; SEM, standard error of mean; TPM, transcripts per million.

We next reclassified all reported PTCs by comparing location with occurrence in ClinVar vs. gnomAD using this NMD prediction, defining classification outcomes based on a ‘confusion matrix’ framework (Fig. 1B). The classical binary model showed little discriminatory power for this task, with no significant separation between ClinVar and gnomAD truncating variants (odds ratio, 0.91; 95% CI, 0.55–1.50; *P* = 0.71). NMDetectiveAI performed better than the binary model (odds ratio, 5.85; 95% CI, 3.96–16.0; *P* = 1.23 × 10^-10^), suggesting an association between predicted NMD escape and truncation pathogenicity that was not captured by the binary model. To quantitatively assess the discriminatory performance of this association, we performed receiver operating characteristic (ROC) analysis (Fig. 1C). The ROC analysis using all exons revealed near-random performance, with the area under the curve (AUC) of 0.52.

However, exclusion of exon 12 or variants beyond residue 1,900 within exon 12 improved performance, increasing the AUC to 0.85 and 0.79, respectively. In addition, improvements were consistently observed across classification metrics upon exclusion of exon 12 entirely or just its distal region (e.g., positive predictive value, 78.2% to 89.7%–91.0%; odds ratio, 5.85 to 19.2–23.4) (Table S1). These data suggest that NMD status correlates with truncation pathogenicity: ClinVar variants were enriched in regions predicted to escape NMD, whereas gnomAD variants were enriched in regions predicted to undergo NMD, but an exception was the distal region of exon 12 (residues 1,900–2,248), where additional mechanisms may limit pathogenicity despite escape from NMD (see below).

### ClinVar but not gnomAD *ASXL3* truncations produce gross protein excess

Because *ASXL3* is preferentially expressed in the brain, we next studied *ASXL3* expression in human neural cells. Human iPSCs were transduced with a lentiviral doxycycline (DOX)-inducible EGFP–Neurogenin 2 (NGN2) cassette and differentiated into induced neurons, hereafter referred to as iNeurons (Fig. S1, A and B). RNA-sequencing (RNA-seq) of wild-type (WT) iNeurons revealed induction of neuronal markers along with suppression of pluripotency and cell-cycle markers over three weeks as expected (Fig. S1C). Notably, *ASXL3* and *ASXL1* expression increased during neuronal induction, whereas *ASXL2* gradually decreased (Fig. 1D). The co-induction of *ASXL1* and *ASXL3* argues against a simple LOF model based on lack of paralogue compensation, since *ASXL* paralogues might be expected to otherwise compensate for loss of *ASXL3*.

To dissect how early versus late *ASXL3* truncations impact protein levels, we quantified endogenous ASXL3 protein levels for representative truncations. Regrettably, high-quality antibodies recognizing ASXL3 do not exist. We therefore introduced an in-frame N-terminus 3×FLAG into the endogenous locus of DOX-inducible NGN2-expressing hESCs (NGN2-hESCs, Fig. S1, D and E). We then introduced representative truncations into the FLAG-tagged haplotype. We used exon 5 to represent ‘early truncations’ from the gnomAD variant-enriched region spanning exons 1–10 (p.N133*, p.R132Qfs*12), and exons 11 and 12 to represent ‘late truncations’ from the ClinVar-enriched region (p.E1006*, p.R1036*, Fig. S1F). NMDetectiveAI predicted NMD for the former and NMD escape for the latter (Fig. 1E). Western blotting for Flag showed WT ASXL3 as a single band at the expected 250 kDa size. Early truncations showed no detectable protein at any molecular weight, consistent with NMD. Both late truncations tested showed proteins at the expected lower molecular weight (110 and 113 kDa, respectively).

Surprisingly, these bands were accumulated >25-fold higher than the full-length protein (Fig. 1, F and G). Furthermore, RNA-seq of iNeurons showed lower *ASXL3* mRNA levels in the early, but not in the late truncation model (Fig. 1H). These results suggest that early truncations (i.e. found in gnomAD) undergo NMD, whereas late truncations (i.e. found in ClinVar) evade NMD. These NMD-evading truncations produce aberrantly stabilized ASXL3 protein, suggesting the presence of a C-terminal degron signal.

### Late but not early *ASXL3* truncations drive epigenetic and transcriptional dysregulation

Previous studies in *ASXL1*-mutant mice and leukemia patients have shown altered chromatin accessibility and gene expression(*8, 14, 15*). To investigate downstream consequences of BRS disease-associated late truncations, we generated NGN2-hiPSCs from skin fibroblasts from two patients harboring truncating variants in exon 11 (p.T398Pfs*10 and p.E1006*) and two in exon 12 (p.R1036* and p.W1609*), covering the major hotspots for pathogenic *ASXL3* variants (Fig. 2A and Fig. S2, A to C), along with four healthy individuals. In addition, we included two WT NGN2-hESC lines, one of which was used for CRISPR engineering. hiPSCs and hESCs were differentiated into iNeurons for RNA-seq and assayed for transposon accessible chromatin (ATAC-seq). Principal component analysis (PCA) of RNA-seq and ATAC-seq datasets revealed a clear separation between WT and BRS iNeurons (Fig. 2, B and E), with marked dysregulation of expression across 1,811 genes (Fig. 2C). We further verified the RNA-seq results by RT-qPCR analysis of representative differentially expressed genes, such as *NEIL1*, *WNT10B*, and *FBLN1* (Fig. 2D). We also found increased chromatin accessibility at transcription start sites (TSSs) in BRS iNeurons compared with WT (Fig. 2F). At representative loci, such as *NEIL1*, BRS iNeurons exhibited increased chromatin accessibility compared with WT (Fig. 2G).

**Fig. 2.**
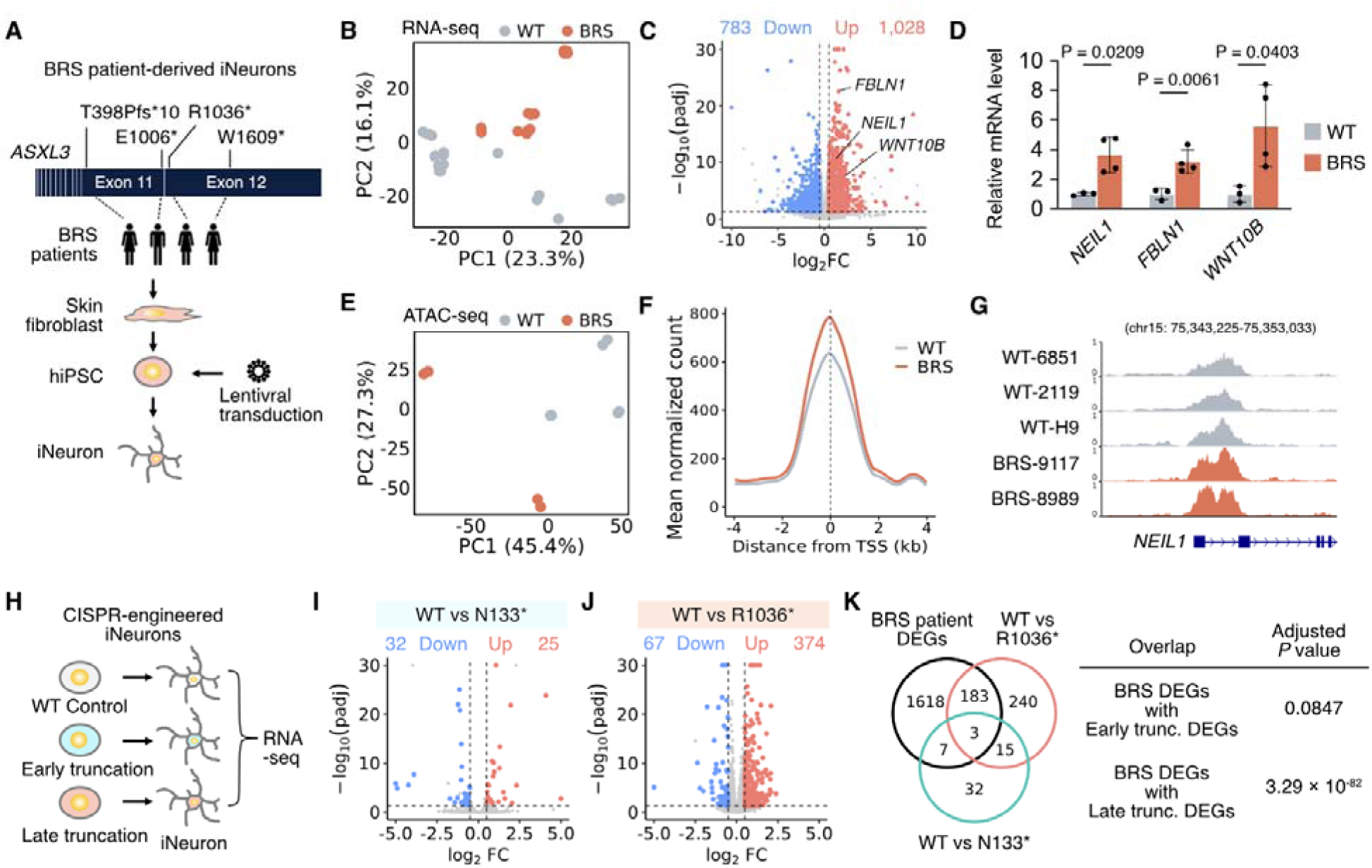
Patient but not control *ASXL3* truncations drive epigenetic and transcriptional dysregulation. (**A**) Generation of BRS patient-derived iNeuron from skin fibroblasts harboring late truncating *ASXL3* variants into iPSCs and differentiated into iNeurons via lentiviral NGN2 transduction. (**B** and **C**) PCA (B) and volcano plot (C) of RNA-seq data from WT (*N* = 6 individuals) and BRS (*N* = 4 individuals) iNeurons at DIV 21. Each individual is represented by *n* = 1–4 biological replicates. In (C), up (red)- or downregulation (blue) were defined by setting *P*adj <0.05 and absolute log2FC ≥ 0.5. Numbers of DEGs indicated. (**D**) Validating the RNA-seq using RT-qPCR for *NEIL1*, *WNT10B*, and *FBLN1*, normalized to *GAPDH*, shown as fold change relative to WT. WT (*N* = 3) and BRS (*N* =4) iNeurons collected at DIV15. Error bars: SEM. Statistical analysis: two-sided Welch’s *t*-test. (**E** and **F**) PCA (E) and Profile plots (F) of ATAC-seq data, showing increased chromatin accessibility around the TSS. WT (*N* = 3 individuals) and BRS (*N* = 2 individuals) iNeurons were collected at DIV 16. Each individual is represented by *n* = 2 biological replicates. In (F), signals averaged across samples within each group. (**G**) ATAC-seq tracks showing increased chromatin accessibility at the *NEIL1* locus. (**H**) RNA-seq from DIV15 iNeurons derived from CRISPR-engineered H9 NGN2-hESC lines. (**I** and **J**) Volcano plots of RNA-seq data comparing WT with p.N133* (I) and p.R1036* (J), showing minimal differential expression in p.N133* and marked dysregulation in p.R1036*. *n* = 3 biological replicates. (**K**) Overlap of DEGs identified in WT vs. BRS iNeurons, H9-WT vs. H9-p.N133* iNeurons, and H9-WT vs. H9-R1036* iNeurons. Venn diagram showing shared DEGs, with strong overlap between BRS and the late-truncation model, but minimal overlap with the early-truncation model. Adjusted *P* values by one-sided Fisher’s exact test with Benjamini–Hochberg correction. Abbreviations: BRS, Bainbridge–Ropers syndrome; PC, principal component; TSS, transcription start sites.

Functional alterations in genes that regulate global gene expression lead to marked transcriptional dysregulation, and the resulting differentially expressed gene (DEG) signature can serve as a disease biomarker(*19*). We therefore sought to leverage our DEG signature as a quantitative readout to compare the transcriptional impact of early and late *ASXL3* truncating variants. We performed RNA-seq using iNeurons differentiated from CRISPR-engineered NGN2-hESCs harboring early (p.N133*) and late (p.R1036*) truncations (Fig. 2H). The early truncation model showed few DEGs, whereas the late truncation model showed marked transcriptional dysregulation (Fig. 2, I and J). In addition, BRS DEGs did not significantly overlap with early truncation DEGs, but showed strong overlap with late truncation DEGs (Fig. 2K), suggesting that an early truncation from the gnomAD-enriched region does not recapitulate the BRS DEG signature. These results suggest a gain-of-function (GOF) effect of late truncating mutations, i.e. with GOF defined as aberrant activity of the truncated protein rather than simply higher protein abundance.

We next hypothesized that, if late truncations act through LOF, forced expression of full-length *ASXL3* might rescue the DEG signature. To test this, we utilized the Sleeping Beauty transposon system to achieve stable expression of FLAG-tagged full-length ASXL3 in CRISPR-engineered NGN2-hESCs (Fig. S3A). We confirmed expression of full-length ASXL3 in both p.N133* and p.R1036* iNeurons, with a consistent increase in *ASXL3* mRNA levels across all lines (Fig. S3, B and C). Forced expression of full-length ASXL3 failed to rescue the DEG signature in either truncation model (Fig. S3, D and E). Rather than reducing the BRS-associated signature, full-length *ASXL3* overexpression only further increased the overlap with BRS DEGs, as might be expected (Fig. S3F). These results suggest that BRS late truncations do not act through a simple LOF mechanism.

### ASO-mediated knock down of *ASXL3* corrects transcriptional GOF in BRS iNeurons

Antisense oligonucleotides (ASOs) enable sequence-specific targeting of gene transcripts and reduce gene expression via RNase H1–mediated degradation. Given the functional redundancy within the ASXL protein family(*20*), we hypothesized that non-allele-selective ASO-mediated knockdown of *ASXL3* could reduce the accumulation of truncated ASXL3 protein and potentially rescue DEG signatures in BRS cells. To test this, we screened 468 candidate 5-10-5 MOE gapmer ASOs targeting distinct sites across the *ASXL3* transcript in SH-SY5Y cells, to identify 14 that achieved >90% knockdown (Fig. S4A). These top-performing ASOs were then assessed in WT iNeurons by evaluating knockdown efficiency 6 days after gymnotic treatment (Fig. 3A). The top candidate (*ASXL3* ASO-4) achieved up to ∼95% reduction relative to the scramble, used for subsequent experiments. (Fig. 3B). We confirmed that this ASO exerts minimal changes on the global transcriptome, likely due to the presence of ASXL1 and 2 (Fig. 3C). Using FLAG-tagged ASXL3 iNeurons, we confirmed that ASO treatment reduced p.R1036* truncated ASXL3 protein levels by ∼90% (Fig. 3, D and E).

**Fig. 3.**
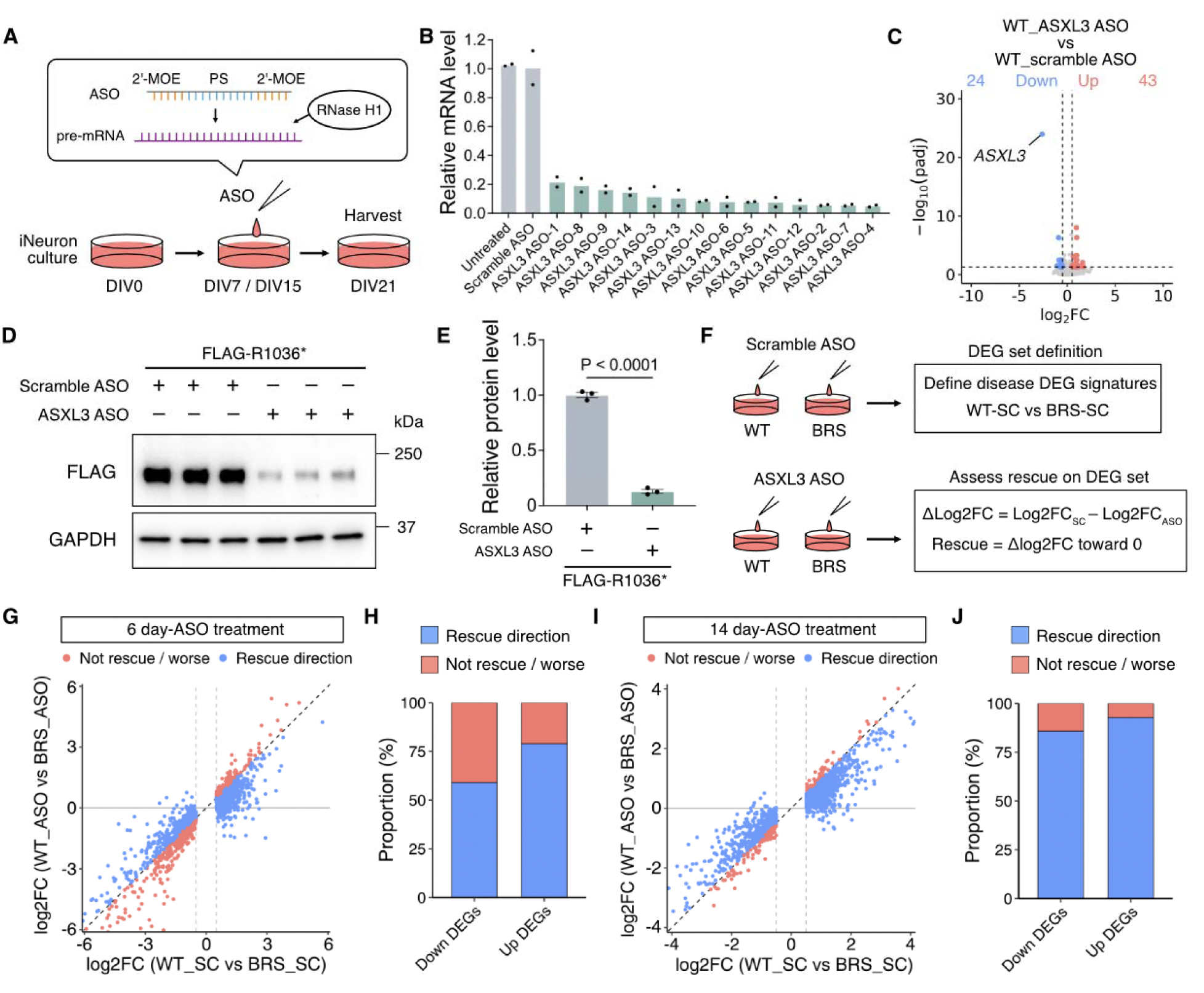
ASO-mediated reduction of ASXL3 protein levels rescues disease DEG signatures. (**A**) 20 nt PS 2’-MOE gapmer ASO and ASO treatment in iNeurons. ASOs hybridize to target pre-mRNA, recruiting RNase H1 to induce site-specific RNA cleavage and degradation. The central DNA gap (blue) is flanked by 2′-MOE–modified bases (red), connected by PS-backbone. (**B**) *ASXL3* mRNA levels measured by RT–qPCR, assessing the knockdown efficiency of candidate ASOs. mRNA expression levels normalized to *GAPDH*. Data shown as fold change relative to scramble ASO-treated controls. *n* = 2 biological replicate. Error bars: SEM. (**C**) Volcano plot of DEGs between WT iNeurons treated with *ASXL3*-targeting ASOs (*N* = 6 individuals) and scramble ASOs (*N* = 6 individuals), from DIV15 to DIV21. Each individual represented by *n* = 1-4 biological replicates. Up (red)- or downregulation (blue) defined by setting *P*adj <0.05 and absolute log2FC ≥ 0.5. Numbers of DEGs indicated. (**D**) Western blot of FLAG-tagged ASXL3 demonstrating efficient knockdown by *ASXL3* ASOs. p.R1036* iNeurons treated with scramble or ASXL3-targeting ASOs, from DIV15 to DIV21. (**E**) Quantification of FLAG-tagged ASXL3 (normalized to GAPDH). Fold change relative to scramble ASO-treated controls. *n* = 3 biological replicates. Error bars: SEM. Statistical analysis: two-sided Welch’s *t*-test. (**F**) ASO efficacy assessed by log2FC values between scramble and *ASXL3* ASO. Rescue: shift toward WT levels (log2FC approaching zero). (**G** and **I**) Scatter plots comparing baseline differential expression (WT_SC vs BRS_SC, x-axis) with *ASXL3* ASO-treated contrast (WT_ASO vs BRS_ASO, y-axis) after 6-day (G) and 14-day. (**I**) ASO treatment. WT (*N* = 6 individuals), BRS (*N* = 4 individuals). Each individual represented by *n* = 1-4 biological replicates. Dashed diagonal line: no change relative to baseline. Genes shifting toward zero highlighted in blue; genes maintained or showing further deviation in red. Axes limited to the 1st-99th percentile to enhance visualization while retaining outliers (31 and 34 data points fall outside the limits in panels G and I, respectively). (**H** and **J**) Proportion of genes showing rescue, or no rescue direction (blue, and red, respectively) among downregulated and upregulated DEGs following 6-day (H) and 14-day (J) ASO treatment, suggesting a more pronounced rescue effect after 14 days and a greater effect in upregulated DEGs than in downregulated DEGs. Abbreviations: PS, phosphorothioate; 2’-MOE, 2’-O-methoxyethyl.

We next asked whether *ASXL3* ASO treatment could rescue DGEs in BRS iNeurons. Compared to scramble ASO-treated conditions, *ASXL3* ASO treatment reduced the number of DEGs from 1,712 to 1,084, indicating a shift toward a more WT-like state (Fig. S4, B and C). We then quantified gene-level rescue by examining changes in the log2 fold change (log2FC) of DEGs defined under scramble ASO conditions (Fig. 3F). We found that after 6 days of treatment, 59.0% (*P* = 2.497 × 10^-5^) of downregulated genes and 78.9% (*P* = 3.831 × 10^-80^) of upregulated genes were rescued (i.e. moved in the direction of wildtype). This effect was further enhanced after 14 days of treatment, reaching 85.7% (*P* = 3.436 × 10^-96^) and 92.7% (*P* = 7.279 × 10^-129^), respectively (Fig. 3, G to J and Fig. S4D). To more quantitatively capture the extent of rescue at the gene level, we defined ‘percent rescue’ for each DEG as the relative shift in log2FC after *ASXL3* ASO treatment. Using this metric, we found that the rescue effect increased with longer treatment: the median percent rescue rose from 29.0% after 6 days of treatment to 37.4% after 14 days of treatment for upregulated genes, and from 4.2% to 26.8% for downregulated genes (Fig. S4E). Together, these results indicate that *ASXL3* ASO treatment restores the BRS transcriptome toward a more WT-like state, likely by reversing *ASXL3* GOF effects.

### ASXL3 C-terminus serves as a ubiquitin-ligase mediated destabilization signal

The marked accumulation of truncated ASXL3, along with its transcriptional GOF effects, suggested that ASXL3 function is tightly coupled to protein abundance. We therefore hypothesized that full-length ASXL3 may undergo rapid proteasomal turnover, resulting in tightly controlled protein abundance. To test this, we generated N-terminally 3×FLAG-tagged full-length ASXL3 and p.R1036* constructs, expressed in HEK293T cells. Treatment with the protein synthesis inhibitor cycloheximide (CHX) showed that full-length ASXL3 levels declined over time, whereas p.R1036* ASXL3 was relatively stable, consistent with impaired protein turnover after C-terminal truncation (Fig. S5, A and B). Treatment with the proteasome inhibitor MG132 increased the abundance of full-length but not p.R1036* ASXL3, suggesting proteasomal degradation of full-length ASXL3 (Fig. S5, C and D).

Proteasomal degradation is largely mediated by the cullin (CUL) family of E3-ligases, so we next sought to identify CUL members responsible for ASXL3 turnover. To this end, we utilized dominant-negative forms of CUL family proteins (DN-CUL), which lack E3 ligase activity but retain the ability to bind substrate recognition modules, thereby blocking ubiquitin-mediated degradation(*21*) (Fig. S5E). Consistent with a previous study demonstrating the interaction between ASXL C-terminus and the CUL4 adaptor DDB1(*22*), co-transfection of DN-CUL4B increased abundance of full-length ASXL3 (Fig. S5, F and G). These results suggest that ASXL3 undergoes ubiquitin–proteasomal degradation in a CUL4-dependent manner.

### PTC position in the last exon dictates ASXL3 protein abundance via a C-terminal destabilization gradient

Based on our observation that numerous very late truncating mutations are present in healthy individuals (after ∼1,900 residues), we hypothesized that not only evasion of NMD but also loss of a C-terminal destabilization signal is required for disease (Fig. 4A). Accordingly, progressively more upstream PTCs would be expected to remove larger portions of this destabilizing region, which may increase truncated protein stability and contribute to greater pathogenicity. To test this, we generated a series of ASXL3 constructs harboring PTCs positioned progressively upstream across exons 11–12, expressed in HEK293T cells (Fig. 4B). Protein accumulation increased stepwise with more upstream truncations, suggesting that ASXL3 C-terminal region is necessary for regulated protein levels (Fig. 4, C and D).

**Fig. 4.**
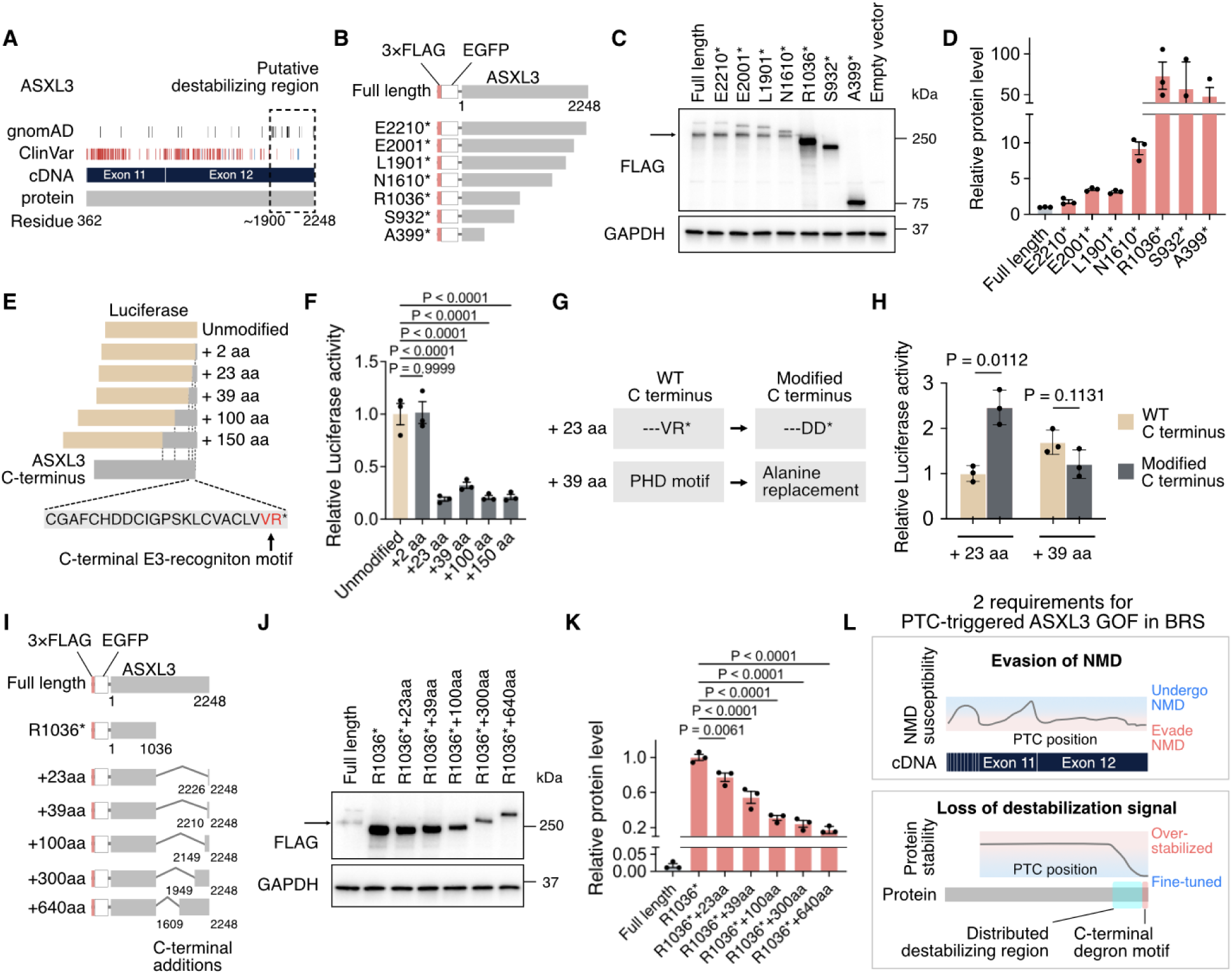
PTC position dictates ASXL3 protein abundance via a C-terminal degradation signal. (**A**) ASXL3 cDNA and protein structure showing the distribution of gnomAD and ClinVar variants. The dashed box indicates a putative destabilizing region located in the C-terminus (∼residues 1,900–2,248). (**B**) Schematic of ASXL3 constructs with progressive C-terminal truncations, which were transfected into HEK293T cells. (**C**) Western blot of FLAG-tagged ASXL3 showing PTC position-dependent protein abundance, increasing with more upstream truncations. GAPDH: protein-loading control. Arrow: nonspecific low-molecular-weight fragment. (**D**) Quantification of FLAG-tagged ASXL3 (normalized to GAPDH). Data shown as fold change relative to full-length ASXL3. *n* = 3 biological replicates. Error bars: SEM. Break in y-axis was used to accommodate all variants. (**E**) Luciferase fusion constructs used to assess degron in ASXL3 C-terminus. The C-terminal VR motif is highlighted in red as an E3 ligase recognition motif. (**F**) Relative luciferase activity of the indicated constructs, showing that the ASXL3 C-terminal 23-aa fragment is sufficient for degron effect. Fold change relative to unmodifiswswwed luciferase. *n* = 3 biological replicates. Error bars: SEM. Statistical analysis: one-way ANOVA followed by Dunnett’s multiple comparisons test. (**G**) Luciferase fusion constructs used to assess the degron effect of the ASXL3 C-terminus. The VR motif was mutated to DD in the +23 aa construct, and the PHD motif within the +39 aa construct was disrupted by alanine substitution. (**H**) Fold change relative to the +23 aa construct with WT C-terminus measured by relative luciferase activity. *n* = 3 biological replicates. Error bars: SEM. Statistical analysis: two-sided Welch’s *t*-test. (**I**) ASXL3 constructs with progressive C-terminal additions to assess the degron effects. (**J**) Western blot of FLAG-tagged ASXL3 showing fragment size-dependent reduction in protein abundance. GAPDH used as control. Arrow: nonspecific low-molecular-weight fragment. (**K**) Quantification of FLAG-tagged ASXL3 (normalized to GAPDH), showing that addition of 640 aa to p.R1036* resulted in ∼85% reduction in protein abundance. Fold change relative to p.R1036* truncated ASXL3. *n* = 3 biological replicates. Error bars: SEM. Break in y-axis was used to accommodate all variants. Statistical analysis: one-way ANOVA with Dunnett’s multiple comparisons test. (**L**) Proposed requirements for disease-causing truncating variants include both escape from NMD as well as removal of C-terminal degron signal. Abbreviations: DD, aspartic acid-aspartic acid; PHD, plant homeodomain; VR, valine–arginine.

We next asked which region of ASXL3 is responsible for its destabilization. A prior study of global protein stability (GPS) using systematic analysis of all possible 23-mer C-terminal sequences found that degradation was promoted if the last two amino acid (aa) residues in a protein were ‘VR’(*23*). Given that the last two residues of all ASXL proteins are ‘VR’, we hypothesized that this C-terminal degron motif regulates ASXL protein abundance. We thus tested sufficiency of ASXL3 C-terminus to promote destabilization, using luciferase fusion constructs (Fig. 4E). We found that, although the VR dipeptide alone did not affect protein abundance, the ASXL3 23-aa C-terminus reduced luciferase activity by 80%, and further extension of the fragment did not enhance this effect (Fig. 4F). We verified that mutation of the VR motif within the 23-aa C-terminus abolished this destabilization effect and significantly increased protein abundance (*P* = 0.0112), suggesting a critical role for the C-terminal VR motif in mediating ASXL3 destabilization (Fig. 4, G and H).

In addition, we noted that all ASXL proteins contain a plant homeodomain (PHD) motif at the C terminus, characterized by the C4HC3 signature. Since some PHD domains have been proposed to have E3 ligase function(*24*), we assessed the potential role of the ASXL3 PHD domain in protein stability by replacing the domain with alanines, within the context of the C-terminal 39-aa residues. We found that this substitution did not affect the reporter protein abundance (Fig. 4, G and H), arguing against a major role of PHD in regulating ASXL3 levels.

To confirm the destabilizing effect of the C-terminus in the context of ASXL3, we added various lengths of the natural C-terminal ASXL3 tail to the truncated p.R1036* ASXL3 by progressively extending C-terminal additions (+23 to +640 aa), expressed in HEK293T cells (Fig. 4I). Surprisingly, the 23-aa fragment conferred only mild reduction of p.R1036* protein accumulation within the ASXL3 context, whereas longer extensions progressively reduced protein levels, with the +640-aa construct showing an ∼85% reduction relative to p.R1036* (Fig. 4, J and K). These results suggest that ASXL3 destabilization is not mediated solely by the context-dependent C-terminal degron motif, but instead involves a broader destabilizing region spanning several hundred residues within the C-terminus, consisting almost entirely of unstructured regions.

We next asked whether specific motifs within the 640-aa region responsible for the ∼85% reduction in p.R1036* abundance. Within the 640-aa region there is a stretch of 25 aa conserved in ASXL paralogues termed the ASXM2 domain, which was also tested for possible degron activity. However, we found that deletion of the ASXM2 domain did not affect ASXL3 abundance. In addition, we tested three additional possible degron motifs within the 640-aa region of ASXL3 predicted by the DegPred program(*25*). However, the deletion of any single of these motifs did not affect protein abundance (Fig. S5, H to J), further supporting that a “broad C-terminal destabilizing region” is required for the regulation of ASXL3 protein levels. These results may explain why very late truncating variants are not disease-causing despite escaping NMD. Overall, our data suggest that the combined effects of NMD escape and loss of a destabilizing signal are critical drivers of ASXL3 GOF underlying BRS (Fig. 4L).

## Discussion

Our data support a ‘two-hit’ model in which NMD escape permits mutant protein production, and loss of the C-terminal degradation signal allows protein accumulation that perturbs transcriptional control. Importantly, this mechanism provides a useful explanation for the variant-position dependence of disease risk among late truncations: those that escape NMD but retain much of the destabilizing C-terminal segment may still be attenuated, whereas truncations that remove the degron-like region can produce the highest protein abundance and the strongest transcriptional effects. In this framework, pathogenicity emerges from the combination of transcript survival and protein stabilization, rather than from coding disruption alone. This principle may extend beyond *ASXL3* to other genes in which distal truncations alter post-translational turnover, helping reconcile why some apparently similar nonsense variants are clinically severe while others are relatively benign. In fact, based upon this GOF prediction, a patient with BRS is currently under study to evaluate the effectiveness of an ASO(*26*).

NMD is a key gatekeeper separating simple LOF from truncation-driven GOF. In our study, the pathogenic *ASXL3* variants were concentrated in positions predicted to escape NMD, allowing production of abundant truncated protein. In contrast, early truncations were largely depleted at the RNA level. This pattern is consistent with broader observations that PTCs near the 3′ end of a gene can evade NMD and, when translated, may generate toxic proteins rather than null alleles(*27*).

For *ASXL3*, NMD escape appears to be a necessary first step, but not sufficient for disease, because very late truncations that escape NMD are seemingly benign. In our data, progressive truncations revealed a stepwise increase in protein accumulation, whereas progressive re-addition of C-terminal fragments correspondingly reduced it, indicating that pathogenicity is not explained solely by NMD escape but also by loss of a destabilization signal within the distal C terminus. This is consistent with the idea that the ASXL3 C terminus contains a built-in proteostatic “brake” that limits steady-state protein levels, so truncations that sever this region convert a normally short-lived chromatin regulator into a stable aberrant product. Our data showed that not only the well-established C-terminal VR dipeptide motif, but also a broader region contributed to degron activity, raising the possibility that the destabilizing signal is distributed across the C-terminal long intrinsically disordered region conserved among ASXL proteins. This model helps explain why some truncations in the 3′ region cause disease while others are tolerated, and it reinforces the idea that variant interpretation should consider both transcript fate and the biochemical behavior of the truncated protein.

A useful way to frame these data is that truncating variants are not mechanistically uniform: in cancer, many recurrent truncations can create stable neomorphic proteins with dominant GOF effects, whereas in neurodevelopmental disorders the same variant class is often assumed to act through haploinsufficiency or complete LOF. Our findings place *ASXL3* squarely in this broader paradigm, by showing that pathogenic late truncations escape NMD, accumulate as aberrantly stabilized proteins, and drive a transcriptional and chromatin-accessibility program that is resistant to simple restoration of full-length protein, consistent with a GOF mechanism. This interpretation aligns with prior work in *ASXL1*-mutant myeloid malignancies, where truncating proteins enhance chromatin dysregulation and promote disease rather than simply reducing dosage(*14, 15*).

In neurodevelopmental disorders, the distinction between LOF and GOF is particularly important because many genes with truncating pathogenic variants are initially annotated as null, even when the protein product can still exert toxic effects(*27–29*). In *ASXL3*, the clinical enrichment of truncations in the NMD-escape region, together with the strong rescue obtained with *ASXL3*-targeting ASOs, suggests that disease depends on both escape from NMD and the preservation of a stable C-terminally truncated protein, and provides proof-of-concept for therapeutic reversibility of the associated molecular phenotype. This model helps reconcile the paradox that early truncations may be tolerated in population databases while late truncations are highly pathogenic, and it supports a framework in which variants in the same gene can produce different consequences depending on where truncations occur and whether the resulting protein gains new properties.

### Study limitations

Although we evaluated several representative human *ASXL3* mutations, we did not evaluate all known mutations for NMD escape, and thus there may be some mutations that do not conform to our models. 2. There is some debate about definition of LOF vs. GOF in the genetics literature. We defined LOF as loss of gene/protein function, and GOF as a state in which a mutant protein acquires aberrant or excessive activity. 3. Although our data support GOF mechanisms, we did not fully exclude potential dominant-negative effects. In particular, we did not directly examine whether the mutant allele interferes with WT protein complexes, or gains new function to disrupt activity. 4. Although many early ASXL3 truncations are evident in healthy individuals, we did not fully exclude the possibility of subtle human phenotypes resulting from early truncations.

## Materials and Methods

### Study design

The objective of this study was to elucidate the functional consequences of *ASXL3* truncating variants identified in healthy individuals and BRS patients, and to determine whether ASO-mediated knockdown can reverse DEG signatures underlying BRS. This study included WT and CRISPR-engineered hESCs as well as hiPSCs derived from healthy individuals and BRS patients that were diagnosed by exome and genome sequencing, and who contributed cells towards generation of iPSCs. The use of hESC/hiPSC lines was approved by the University of California San Diego Institutional Review Board (#147028). *ASXL3*-targeting ASOs were first screened in SH-SY5Y cells, followed by treatment in WT iNeurons to confirm their efficacy. ASO treatment of iNeurons was performed using gymnotic uptake, which is predictive of in vivo uptake. No statistical analysis was done to calculate sample size. No specific rules for the stopping of data collection were made. All data collected were included in the study. All experiments were done with a minimum of three biological replicates, unless otherwise specified. No outliers were removed from the data.

### Correlation between *ASXL3* truncating variants and predicted NMD escape

Truncating variants were obtained from ClinVar (accessed 31 March 2026; filtered to include frameshift and nonsense variants, *n* = 220) and gnomAD (v4.1.0; filtered to include frameshift and stop gained variants, *n* = 90). NMDetectiveAI was used to assign a predictive NMD efficiency score to each PTC(*30*). To define a score for which higher values correspond to stronger predicted NMD escape, we derived an NMD escape score as −1× the NMD efficiency score. For visualization, NMDetectiveAI prediction scores across ASXL3 PTC positions were transformed to z-scores and displayed as a heatmap to highlight relative positional trends in predicted NMD susceptibility. A ROC curve and corresponding AUC were generated using the pROC R package(*31*), treating ClinVar PTCs as the positive (disease associated) class and gnomAD PTCs as the negative (tolerated) class. The optimal NMD escape score threshold was chosen using Youden’s index, to maximize the sum of sensitivity and specificity. This threshold was then used to dichotomize the NMD escape score, and 2×2 contingency tables constructed at this cutoff were analyzed using two-sided chi square tests to assess statistical significance.

### Cell culture

A summary of hESC and hiPSC lines used is listed in Table S2. hESCs and hiPSCs were cultured in mTeSR plus media (STEMCELL Technologies, #100-0276) on hESC-Qualified Matrigel coated dishes (Corning, #354277). HEK293T cells were cultured in DMEM supplemented with 10% FBS and 1% Pen/Strep. Human SH-SY5Y neuroblastoma cells were maintained in DMEM/F-12 (Thermo Fisher Scientific, #10565018) supplemented with 10% FBS and 1% Pen/Strep. For differentiation, SH-SY5Y cells were seeded onto pre-coated poly-D-lysine 96-well tissue culture plates (Corning, 356461), then 1d later, growth medium was replaced with “differentiation medium” consisting of Neurobasal (Thermo Fisher Scientific, #21103049) supplemented with 1× B-27 (Thermo Fisher Scientific, #0080085SA), 1×GlutaMAX and 10 µM all-trans retinoic acid (Sigma-Aldrich, #R2625), plus 1% Pen/Strep. Cells were differentiated for 10d, with half-volume medium changes every 2–3d. All active cultures tested negative monthly for mycoplasma.

### Plasmid construction

Plasmids were modified by site-directed mutagenesis or NEBuilder HiFi DNA assembly (New England Biolabs, #M0494S, #E2621S). An N-terminal 3×FLAG-EGFP tag was inserted into the pcDNA3.1 full-length ASXL3 construct (gift from S. Bielas). All constructs verified by whole-plasmid sequencing (Plasmidsaurus).

### iPSC-derived neurons

Lentiviral transduction and neuronal differentiation were performed as previously described(*32*). In brief, lentivirus was generated by transfecting HEK293T cells with pMD2.G (Addgene, #12259), psPAX2 (Addgene, #12260), and pLVX-UBC-rtTA-NGN2:2A:EGFP (Addgene, #127288) plasmids. hESCs and hiPSCs were transduced with lentivirus and subsequently selected with 0.5 μg/ml of puromycin (Sigma-Aldrich, #127288). Induction was initiated when selected colonies reached 60-70% confluence by adding 2 µg/mL DOX (Sigma-Aldrich, #D3447). On DIV2, the culture medium was switched to neuronal medium containing 1:1 ratio of DMEM/F-12 and Neurobasal, 1× N2 supplement (Thermo Fisher Scientific, #16502048), 1× B27 supplement, 1× NEAA (Thermo Fisher Scientific, #11140050), 1ug/ml laminin (Sigma-Aldrich, #A29248), and 2ug/ml DOX. On DIV3, cells were passaged to the plates coated with 30 μg/mL poly-l-ornithine (Sigma-Aldrich, #P4957) and 4 μg/ml laminin. On DIV5-7, proliferating cells were eliminated by treatment with 4 μM cytarabine (Sigma-Aldrich, #1162002) for 48h. Thereafter, the culture medium was replaced every 3d until harvest.

### Generation of CRISPR-engineered NGN2-hESC lines

CRISPR-mediated knock-in was carried out as previously described(*33*). In brief, Cas9 ribonucleoprotein (RNP) complexes were assembled using Alt-R crRNA and ATTO-550-labeled tracrRNA (Integrated DNA Technologies) and delivered into hESCs together with a GFP plasmid and single-stranded oligodeoxynucleotide (ssODN) donor by electroporation (Nucleofector 2b, Lonza; program B-016). Cells were cultured with 0.5 µM HDR enhancer V2 for 24h, and ATTO-550⁺/GFP⁺ double-positive cells were isolated by FACS at 48h. Early truncating variants were introduced via the non-homologous end-joining (NHEJ) pathway without the use of an ssODN donor and an HDR enhancer. Sorted cells were plated at low density for clonal isolation, and individual clones were screened by Sanger sequencing or Oxford Nanopore sequencing of the targeted locus. All oligos are listed in Table S3.

### Western blotting

Cells were washed 3× with DPBS and lysed with radioimmunoprecipitation buffer (RIPA, 10 mM Tris-HCl, pH 8.0, 1 mM EDTA, 0.5 mM EGTA, 1% Triton X-100, 0.1% sodium deoxycholate, 0.1% SDS and 140 mM NaCl) supplemented with protease inhibitor cocktail (Sigma-Aldrich, #11697498001). Lysates were centrifuged at 20,000g for 10 min at 4 °C. The supernatant was collected for BCA assay (Thermo Fisher Scientific, #23225) and mixed with 4× SDS sample buffer containing β-mercaptoethanol. 10 μg of total lysates were separated by SDS-PAGE and analyzed by western blotting. Images were captured using iBright FL1500 (Invitrogen). Quantification of protein band intensity was performed using the ImageJ band densitometry plug-in. Primary antibodies used include mouse anti-FLAG M2 (Sigma-Aldrich, #F1804) and mouse anti-GAPDH (Abcam, #8245).

### qPCR

Cells were lysed in TRIzol (Invitrogen, #15596026) according to manufacturer instructions. A total of 500 ng RNA was used for cDNA synthesis with the SuperScript III First-Strand Synthesis System (Invitrogen, #18080051). RT–qPCR was performed using SYBR Green master mix on a CFX Real-Time PCR System (Bio-Rad), and relative mRNA expression was quantified using the 2-ΔΔct method according to the MIQE guidelines(*34*). All primer sequences are listed in Table S3.

### RNA-seq

RNA-seq was performed as previously described(*32*). In brief, libraries were prepared using the TruSeq Stranded mRNA Library Kit (Illumina, #20020595) and sequenced to a depth of at least 10 million reads per sample on an Illumina NovaSeq S4 platform. Raw reads were quality-checked using FastQC, and adapter trimming was performed using Cutadapt. Trimmed reads were aligned to the hg38 reference genome using STAR. Gene-level read counts were quantified using featureCounts, and TPM was estimated using RSEM. Differential gene expression analysis was performed using DESeq2. DEGs were defined as those with an adjusted *P* value < 0.05 and an absolute log2FC > 0.5. For PCA, variance-stabilized counts were calculated using DESeq2.

### ATAC-seq

ATAC-seq was performed on 100,000 DAPI-negative live cells sorted by FACS. Libraries were prepared using the ATAC-seq kit (Active Motif, #53150). Nuclei were isolated and subjected to transposition using a Tn5 transposase, followed by purification and PCR amplification. Indexed libraries were pooled and sequenced on an Illumina NovaSeq X Plus platform using 2×50 bp paired-end reads. Raw reads were processed using the nf-core/atacseq pipeline, including adapter trimming, quality filtering, alignment to the hg38 reference genome with BWA, and removal of PCR duplicates and mitochondrial reads. Peaks were called for each sample using MACS2, and a consensus peak set was generated by merging peaks across all samples. Read counts across the consensus peak set were quantified using featureCounts. Chromatin accessibility was assessed using DESeq2, and differentially accessible regions (DARs) were defined as peaks with an adjusted *P* value < 0.05 and an absolute log2FC > 0.5. For PCA, variance-stabilized counts were calculated without collapsing replicates, and the top 10,000 most variable peaks were used. For visualization of chromatin accessibility around TSS, peak centers were assigned to the nearest TSS based on hg38 annotations, and DESeq2-normalized counts were aggregated within a ±4 kb window.

### *ASXL3* transposition

For stable expression of full-length ASXL3, the Sleeping Beauty transposon plasmid (Addgene, #60516) carrying the 3×FLAG-ASXL3 transgene was electroporated together with a transposase plasmid (Addgene, #34879) into WT, p.N133*, and p.R1036* NGN2-hESC lines, selected for 2w with hygromycin (Sigma-Aldrich, #H3274), induced to neuronal differentiation, then harvested at DIV15.

### ASO design

The ASOs used in this study were designed and synthesized as previously described by Ionis Pharmaceuticals through collaborative work with the n-Lorem Foundation(*35, 36*). In brief, the ASOs were designed as 20-nucleotide gapmers containing a 10-nucleotide phosphorothioate and phosphodiester (PS/PO) mixed backbone deoxynucleotide gap flanked by five 2′-O-methoxyethyl (MOE)-modified nucleotides on both the 5′ and 3′ sides. 468 ASOs were designed and screened in SH-SY5Y cells using standard design algorithms. Following the primary screen, 14 non-allele-selective *ASXL3*-targeting ASO sequences were prioritized based upon achieving >90% knockdown of target RNA, with favorable dose-response characteristics. The non-targeting scramble ASO that has no natural target in human cells was previously published(*37*). All ASOs used in this study are listed in Table S3.

### ASO treatment of iNeurons

ASOs were used at final concentration of 10 µM with gymnotic uptake, incubated for 6 or 14 days before harvesting, with ASOs replenished every 3d. The scramble ASO was used as a negative control at the same concentration. Disease-associated DEGs were defined based on WT versus BRS differences under scramble ASO conditions, and rescue was defined as a shift in log2FC toward zero following *ASXL3* ASO treatment. Percent rescue was calculated as 100 × (baseline log2FC − treated log2FC) / |baseline log2FC| for upregulated DEGs and 100 × (treated log2FC − baseline log2FC) / |baseline log2FC| for downregulated DEGs.

### Transfection

HEK293T cells were seeded and transfected next day with indicated plasmids using Lipofectamine 2000 (Thermo Fisher Scientific, #11668027), then harvested 24h later. MG132 (Sigma-Aldrich, #474790) treatment was initiated at 24h, harvested 12h later.

### Luciferase assay

HEK293T cells were seeded into 96-well plates and co-transfected next day with the modified firefly luciferase plasmids (Addgene, #163523) along with renilla (Addgene, #12179), cells were harvested 24h later, and luciferase activity was measured using the dual-luciferase reporter assay system (Promega, #E1910) on a microplate luminometer (Tecan).

### Statistical analysis

Unless specifically stated, each experiment was performed at least twice for each condition. Statistical analyses were performed with R or Prism v10 (GraphPad). Statistical tests are noted in figure legends.

## List of Supplementary Materials

Fig S1 to S5

Tables S1 to S3

Data files S1 to S4

## Data Availability

RNA-seq and ATAC-seq data are available from the SRA (accession number XXXXX). All study reagents are available upon request to J.G.G., according to the terms of the standard Uniform Biological Material Transfer Agreement.

## Acknowledgements

Authors would like to thank A. Rao and Z. Dong (La Jolla Institute for Immunology), A. Shilatifard (Northwestern U), Y. Chen (UC San Diego), A. Economides (Regeneron) for discussions, S. Bielas and E. Peirent (U Michigan) for discussions and materials, the n-Lorem Foundation and Ionis Pharmaceuticals for sharing data, the Stem Cell Genomics Core at the Sanford Consortium for use of the ARIA FACS instrument. This work was supported by grants from the California Institute for Regenerative Medicine (DISC2-13469), the Simons Foundation for Autism Research Initiative (SFARI-882181), Japan Society for the Promotion of Science Overseas Research Fellowships (202460474), the California Institute for Regenerative Medicine Bridges to Stem Cell Research Internship Program, and Lowy Medical Research Initiative.

## Ethics statement

Human subjects research was conducted in accordance with protocols approved by the University of California San Diego Human Research Protections Program (HRPP)/Institutional Review Board. Written informed consent was obtained from all participants, or from a parent or legal guardian when applicable, using IRB-approved consent forms that included permission for research participation, genetic analysis, and assessment of cellular phenotypes in patient-derived specimens and cellular models. The UC San Diego HRPP operates under Institutional Assurance number FWA00004495.

## Author contributions

Y.N. and T.N. planned and conducted experiments involving tissue culture, data analysis, and writing of the manuscript. N.M. and D.D. aided in acquiring patient-derived iPSC lines. Y.N. and I.T. conducted experiments involving viral transduction. Y.N., T.N., C.J.T., H.T., and R.Y.Z. conducted experiments involving CRISPR genome editing, plasmid construction, and biochemical analyses. W.Z. conducted bioinformatic analyses. K.S.-S., J.D., L.M., A.D., S.F., and A.W. designed, synthesized, and screened ASOs. S.J., S.T.K., and J.G.G. contributed to planning, guiding, and writing the manuscript.

---

**Fig. S1.**
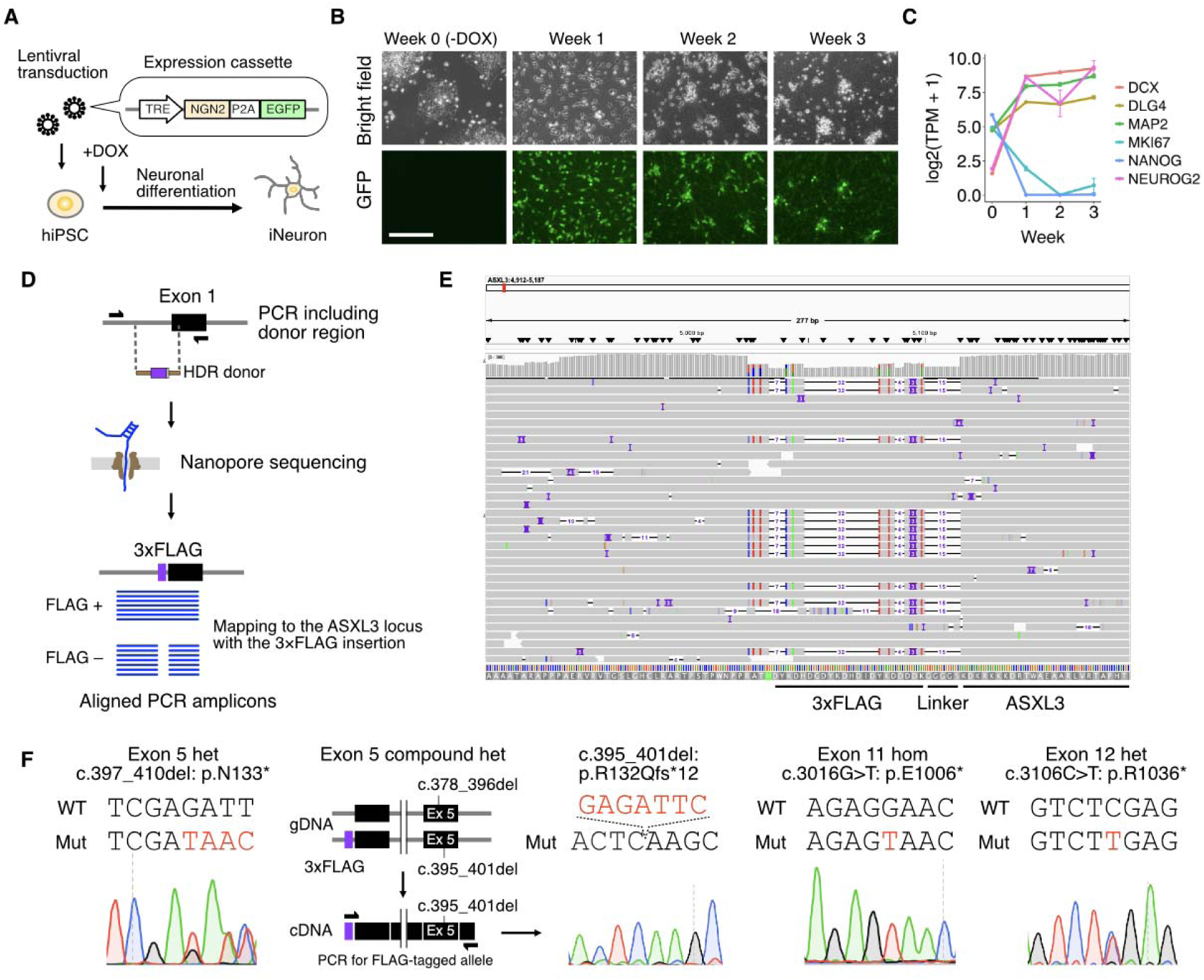
Validation of iNeuron differentiation and genotyping of CRISPR-engineered *ASXL3* mutant lines. (**A**) Generation of iNeurons. hiPSCs transduced with lentivirus containing an NGN2-P2A-EGFP expression cassette. DOX activates NGN2 expression through the TRE promoter, driving neuronal differentiation. (**B**) Representative bright-field (top) and GFP fluorescence (bottom) images of iNeurons at week 0 (i.e. iPSCs prior to DOX treatment),1,2 and 3. Scale bar: 200 µm. (**C**) Temporal expression of neuronal and pluripotency markers during differentiation in WT iNeurons (*n* = 4 biological replicates). Error bars: SEM. (**D**) Schematic illustrating the validation of the N-terminal 3×FLAG knock-in at the endogenous ASXL3 locus. PCR amplification across the donor region followed by nanopore sequencing was used to confirm correct insertion. (**E**) Integrative Genomics Viewer (IGV) view showing heterozygous insertion of the 3×FLAG tag at the endogenous ASXL3 locus. 3×FLAG, linker, and ASXL3 coding regions annotated. Start codon: green. (**F**) Sanger sequencing confirms CRISPR-mediated editing of *ASXL3* in FLAG (+) H9 NGN2-hESCs. Altered sequences: red. Allele-specific RT-PCR confirmed exon 5 c.395_401del mutation on the FLAG-tagged haplotype. Abbreviations: DOX, doxycycline; P2A, 2A self-cleaving peptide; TRE, tetracycline-responsive element.

**Fig. S2.**
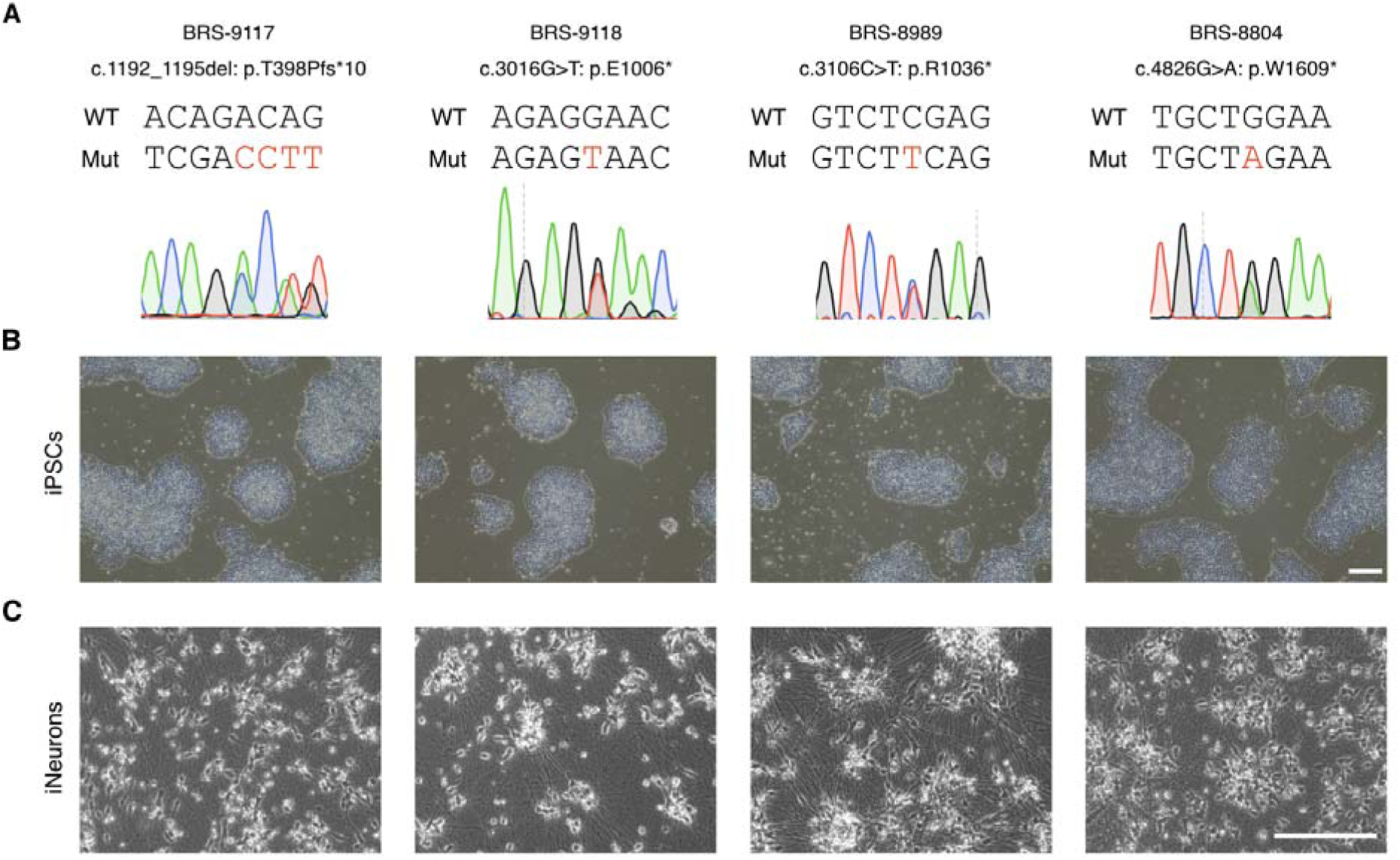
Validation of iNeuron differentiation and genotyping of BRS patient-derived iPSC lines. (**A**) Sanger sequencing of gDNA extracted from BRS patient-derived iPSCs. Altered sequences in red. (**B** and **C**) Representative images of BRS patient-derived iPSCs (B) and iNeurons (C). Scale bar: 200 µm.

**Fig. S3.**
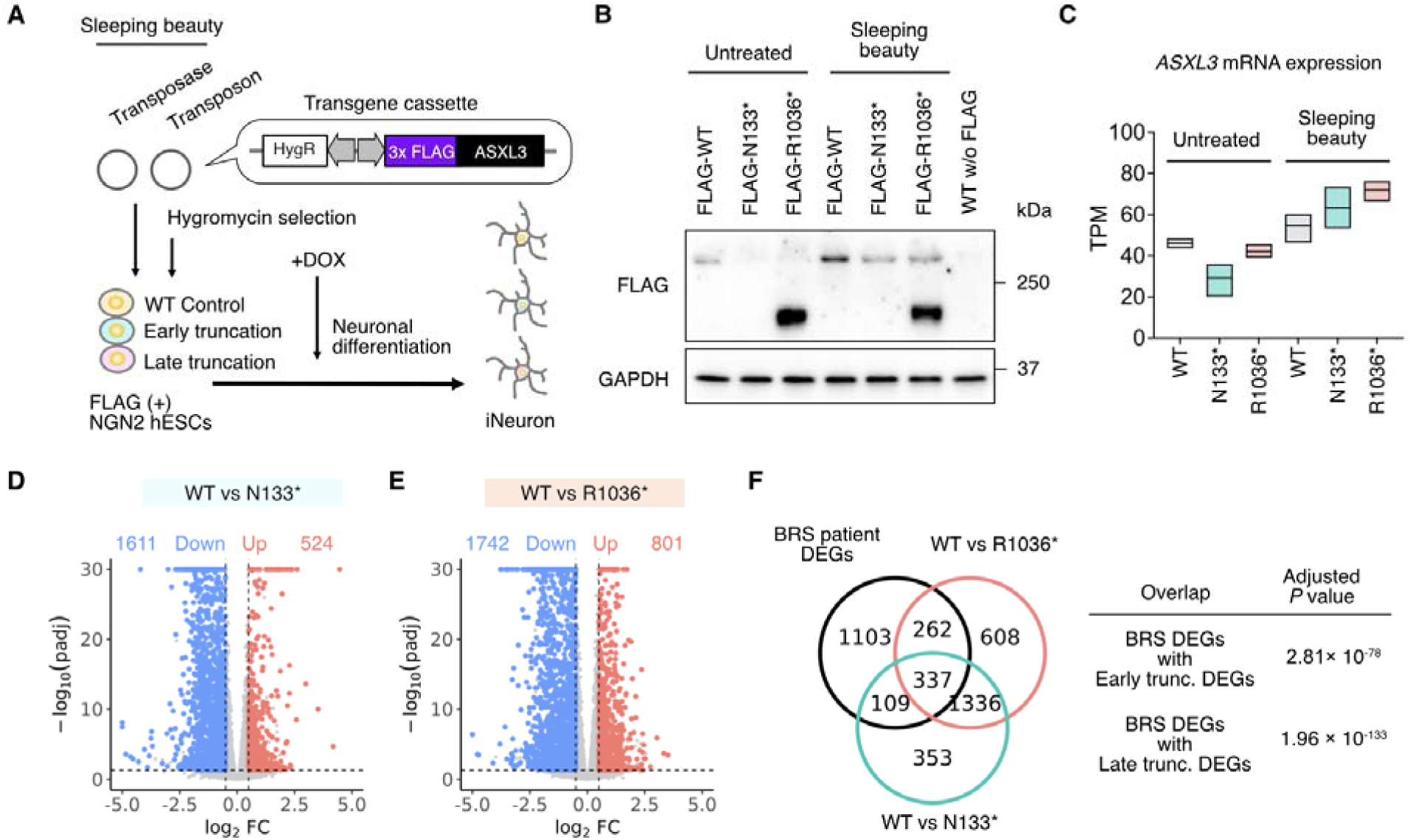
Forced expression of full-length ASXL3 exacerbates transcriptional dysregulation in both early and late truncation models. (**A**) Full-length *ASXL3* constructs stably introduced into NGN2 hESCs harboring FLAG-tagged WT, early truncation, or late truncation *ASXL3* backgrounds using the Sleeping Beauty transposon system, followed by hygromycin selection and DOX-induced differentiation into iNeurons. (**B**) Western blot of FLAG-tagged ASXL3 showing expression of full-length ASXL3 in the Sleeping Beauty-treated group. GAPDH: loading control. Arrow: nonspecific band. (**C**) RNA-seq of DIV15 iNeurons, showing increased *ASXL3* mRNA expression after Sleeping Beauty-mediated forced expression across all genotypes. *n* = 3 biological replicates. Floating bars (min to max). Mean: horizontal line. (**D** and **E**) Volcano plots of RNA-seq data comparing WT with p.N133* (D) and p.R1036* (E) following Sleeping Beauty-mediated forced expression of full-length ASXL3, showing marked transcriptional dysregulation in both groups. *n* = 3 biological replicates. (**F**) Overlap of DEGs identified in WT vs. BRS iNeurons, H9-WT vs. H9-p.N133* iNeurons, and H9-WT vs. H9-R1036* iNeurons. Venn diagram showing shared DEGs, with strong overlap between BRS and both late and early-truncation models. Adjusted *P* values: one-sided Fisher’s exact test with Benjamini–Hochberg correction.

**Fig. S4.**
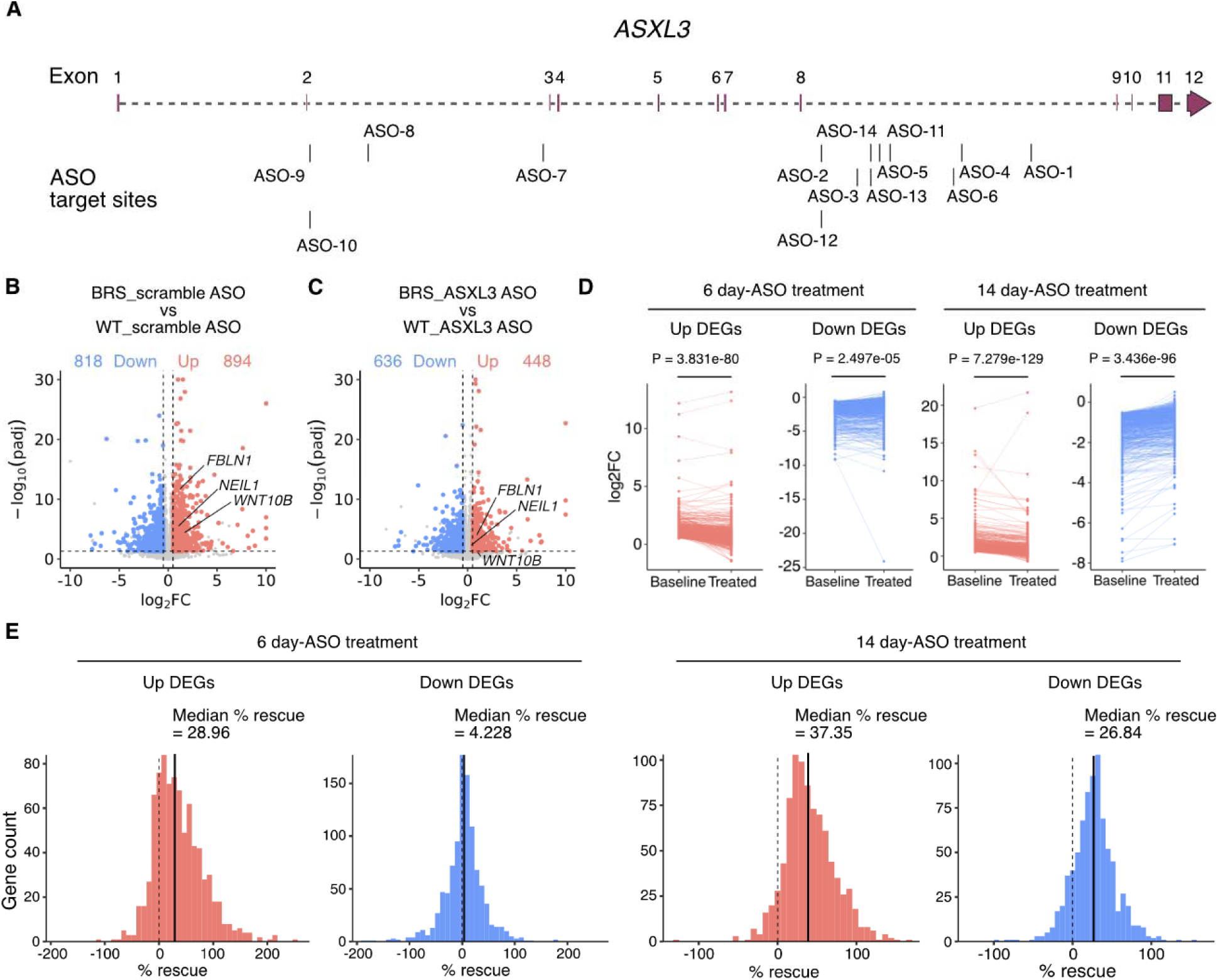
Rescue of disease DEG signatures by *ASXL3*-targeting ASOs. (**A**) Schematic representation of the *ASXL3* gene structure and the positions of ASO target sites. (**B** and **C**) Volcano plots of DEGs, showing fewer DEGs and a shift of *FBLN1*, *NEIL1*, and *WNT10B* toward normalization following *ASXL3* ASO treatment. (B) BRS + scramble ASO (DIV7–DIV21) vs WT + scramble ASO. (C) BRS + *ASXL3* ASO (DIV7–DIV21) vs WT + *ASXL3* ASO. WT (*N* = 6 individuals), BRS (*N* = 4 individuals). Each individual is represented by *n* = 1-4 biological replicates. Up (red)- or downregulation (blue) were defined by setting *P*adj <0.05 and absolute log2FC ≥ 0.5. Numbers of DEGs indicated. (**D**) Gene-wise paired comparison of log2FC between baseline and indicated ASO-treated conditions for upregulated and downregulated DEGs following ASO treatment. Each line represents an individual gene. Statistical analysis: one-sided paired Wilcoxon signed-rank tests. (**E**) Distributions of percent rescue for upregulated and downregulated DEGs following ASO treatment. Each bar represents the number of genes within a given percent rescue interval (40 bins spanning the full range of values). Dashed vertical line at 0%: no rescue. Solid vertical line: median percent rescue across genes.

**Fig. S5.**
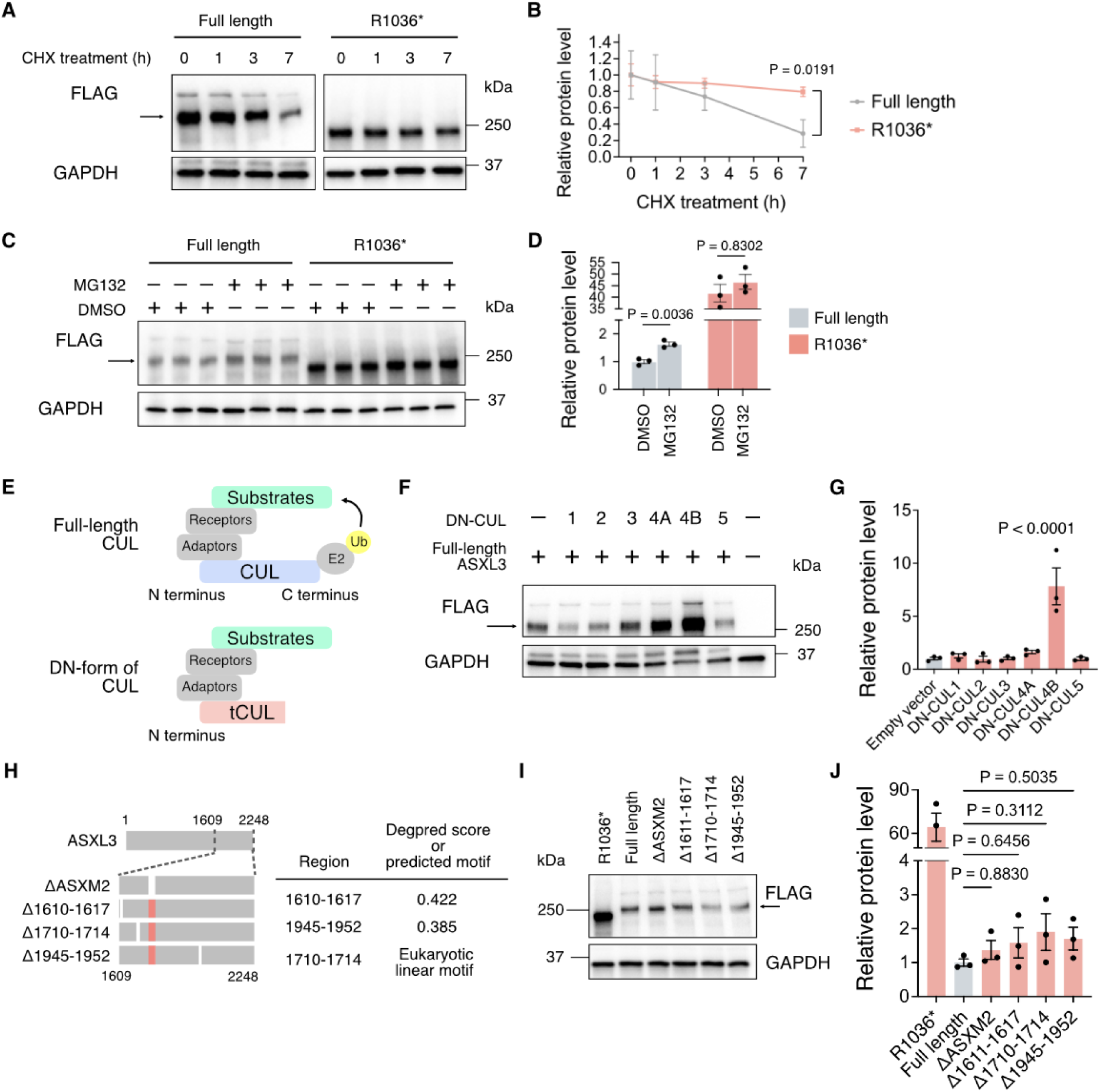
Analysis of ASXL3 protein stability and degradation mechanisms. (**A**) Western blot of FLAG-tagged ASXL3 levels in HEK293T cells expressing the indicated constructs after 20 µg/mL CHX treatment for the indicated times. GAPDH: protein-loading control. Arrow: nonspecific low-molecular-weight fragment. (**B**) Quantification of FLAG-tagged ASXL3 (normalized to GAPDH). Data shown as fold change relative to the 0-h time point for each construct. *n* = 3 biological replicates. Error bars: SEM. Statistical analysis: ordinary two-way ANOVA followed by Šidák’s multiple-comparisons test. (**C**) Western blot of FLAG-tagged ASXL3 levels in HEK293T cells expressing the indicated constructs and treated with DMSO or 20 µM MG132. GAPDH: protein-loading control. Arrow: nonspecific low-molecular-weight fragment. (**D**) Quantification of FLAG-tagged ASXL3 (normalized to GAPDH). Data shown as fold change relative to full-length ASXL3 treated with DMSO. *n* = 3 biological replicates. Error bars: SEM. Statistical analysis: two-sided Welch’s *t*-test. Break in y-axis was used to accommodate all variants. (**E**) Schematic of full-length and DN-CUL proteins. The DN-truncated form (tCUL) lacks the C-terminal domain required for ubiquitination but retains adaptor and receptor binding, thereby inhibiting CUL-dependent ubiquitination. (**F**) Western blot of FLAG-tagged ASXL3 levels in HEK293T cells co-expressing the full-length ASXL3 with the indicated DN-CUL constructs or an empty vector control. GAPDH: protein-loading control. Arrow: nonspecific low-molecular-weight fragment. (**G**) Quantification of FLAG-tagged ASXL3 (normalized to GAPDH). Data shown as fold change relative to the empty vector control. *n* = 3 biological replicates. Error bars: SEM. Statistical analysis: one-way ANOVA with Dunnett’s multiple comparisons test. Only statistically significant comparisons are indicated. (**H**) ASXL3 C-terminal deletions targeting structured domain or predicted degron motifs, including sites with high DegPred scores and a predicted eukaryotic linear motif. (**I**) Western blot analysis of FLAG-tagged ASXL3 levels in HEK293T cells expressing the indicated constructs. GAPDH: protein-loading control. Arrow: nonspecific low-molecular-weight fragment. (**J**) Quantification of FLAG-tagged (normalized to GAPDH). Data shown as fold change relative to full-length ASXL3. *n* = 3 biological replicates. Error bars: SEM. Break in y-axis used to accommodate all variants. One-way ANOVA with Dunnett’s multiple comparisons test. Break in y-axis used to accommodate all variants. Abbreviations: CHX, cycloheximide; DN, dominant negative.

## Supplementary tables

**Table S1.** Region-dependent performance of NMD-based classification of *ASXL3* truncating variants.

| Prediction method | Region analyzed | Positive predictive value (%) | Negative predictive value (%) | Odds ratio (95% CI) | P value |
| --- | --- | --- | --- | --- | --- |
| NMDetectiveAI | Exon 1-11 | 91 | 70.5 | 23.4 (9.22-59.3) | $2.58 \times 10^{-14}$ |
| NMDetectiveAI | Exon 1-12 (residues 1-1,900) | 89.7 | 68.9 | 19.2 (8.90-41.5) | $1.00 \times 10^{-18}$ |
| NMDetectiveAI | Exon 1-12 (residues 1-2,248) | 78.2 | 68.9 | 5.85 (3.96-16.0) | $1.23 \times 10^{-10}$ |
| Classical binary model | Exon 1-12 (residues 1-2,248) | NA | NA | 0.91 (0.55-1.50) | 0.71 |

**Table S2.** Summary of human ESCs and iPSCs.

| Type | ID | Sex | Genotype (NM_030632.3) | Source |
| --- | --- | --- | --- | --- |
| hESC | H1 (WA01) | M | WT | WiCell RRID:CVCL_D092 |
| hESC | H9 (WA09) | F | WT | WiCell RRID:CVCL_9773 |
| hiPSC | WT-6851 | F | WT | WiCell GM27160 S033751B |
| hiPSC | WT-2920 | F | WT | California Institute for Regenerative Medicine CW60490 |
| hiPSC | WT-2116 | M | WT | California Institute for Regenerative Medicine CW60278 |
| hiPSC | WT-2631 | M | WT | California Institute for Regenerative Medicine CW60394 |
| hiPSC | BRS-9117 | F | c.1192_1195del: p.T398Pfs*10 | Simons Foundation Autism Research Initiative 16179-SV0002503 |
| hiPSC | BRS-9118 | M | c.3016G>T: p.E1006* | Simons Foundation Autism Research Initiative 16779-SV0003078 |
| hiPSC | BRS-8989 | F | c.3106C>T: p.R1036* | Simons Foundation Autism Research Initiative 16243-SV0003980 |
| hiPSC | BRS-8804 | F | c.4826G>A: p.W1609* | Gift from Dr. Mor, Sheba Medical Center |

**Table S3:**
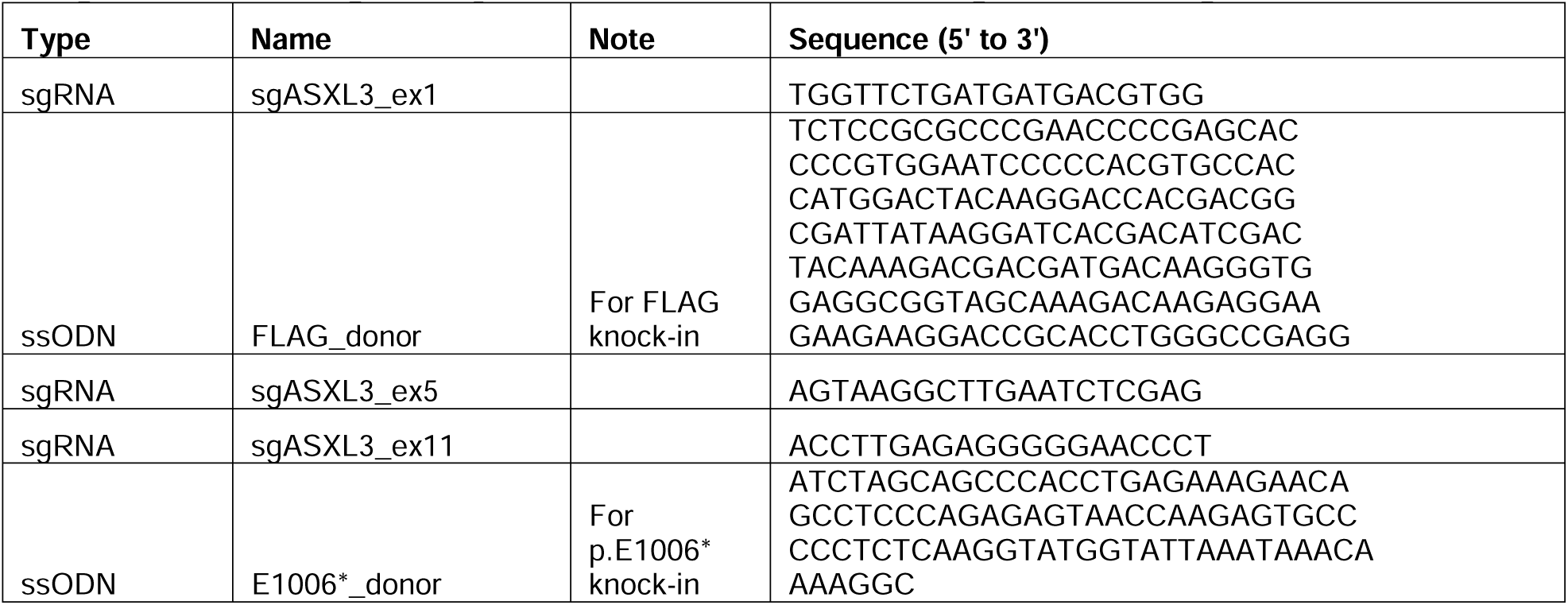
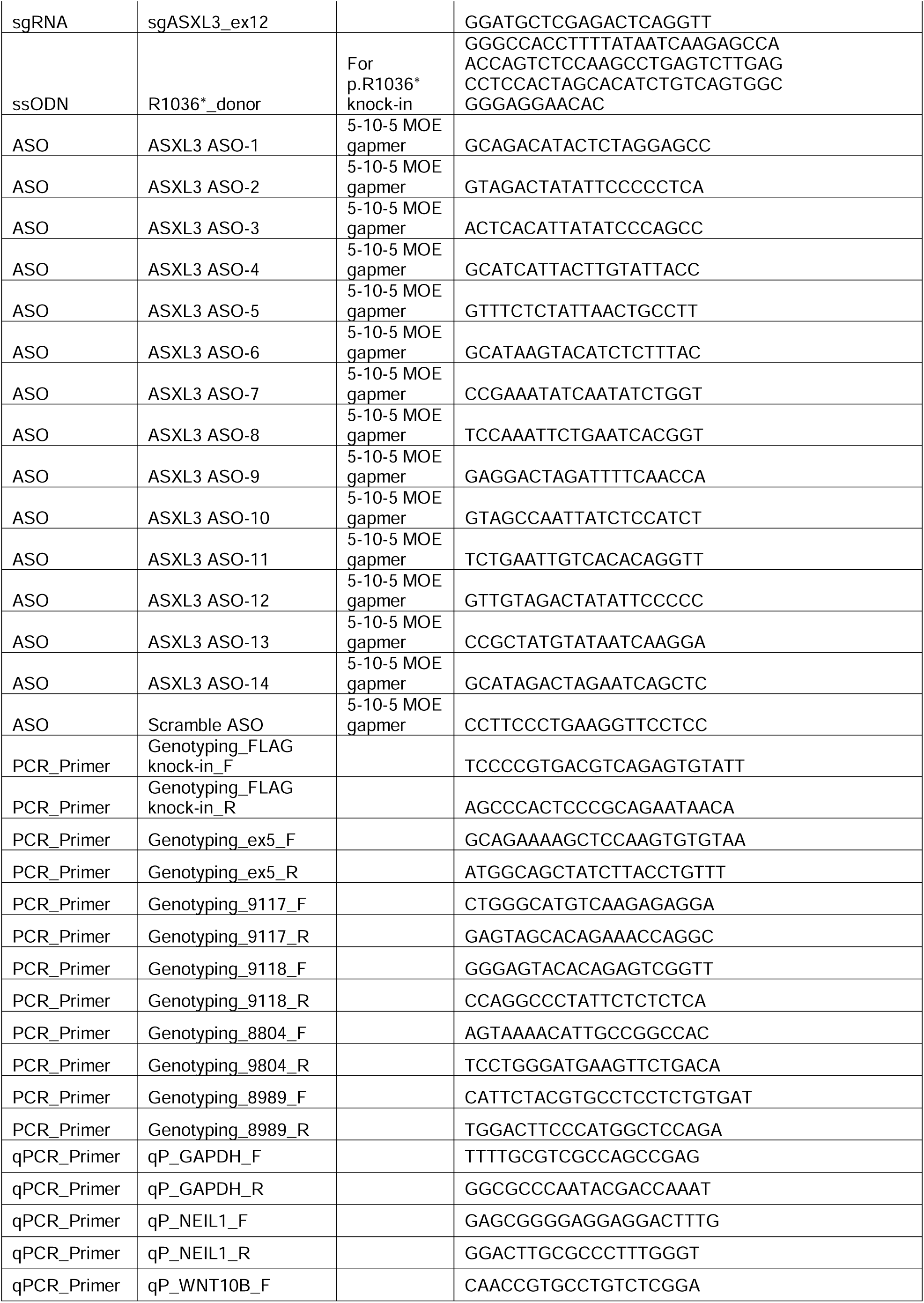
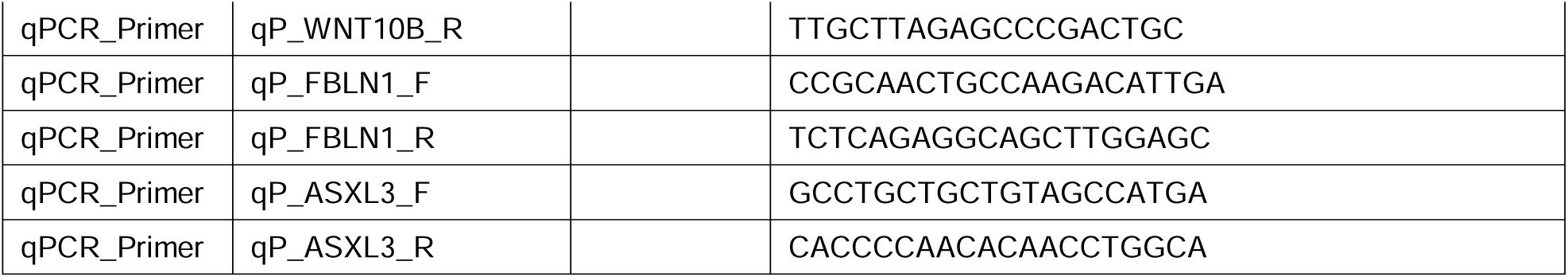
Sequences of primers, oligo nucleotides, antisense oligo nucleotides (ASOs), single-stranded oligodeoxynucleotides (ssODNs), and guide RNAs (gRNAs).

## References and Notes

1. J. C. Scheuermann, A. G. de Ayala Alonso, K. Oktaba, N. Ly-Hartig, R. K. McGinty, S. Fraterman, M. Wilm, T. W. Muir, J. Müller, Histone H2A deubiquitinase activity of the Polycomb repressive complex PR-DUB. Nature 465, 243–247 (2010).

2. O. Abdel-Wahab, M. Adli, L. M. LaFave, J. Gao, T. Hricik, A. H. Shih, S. Pandey, J. P. Patel, Y. R. Chung, R. Koche, F. Perna, X. Zhao, J. E. Taylor, C. Y. Park, M. Carroll, A. Melnick, S. D. Nimer, J. D. Jaffe, I. Aifantis, B. E. Bernstein, R. L. Levine, ASXL1 mutations promote myeloid transformation through loss of PRC2-mediated gene repression. Cancer Cell 22, 180–193 (2012).

3. L. Fagerberg, B. M. Hallström, P. Oksvold, C. Kampf, D. Djureinovic, J. Odeberg, M. Habuka, S. Tahmasebpoor, A. Danielsson, K. Edlund, A. Asplund, E. Sjöstedt, E. Lundberg, C. A.-K. Szigyarto, M. Skogs, J. O. Takanen, H. Berling, H. Tegel, J. Mulder, P. Nilsson, J. M. Schwenk, C. Lindskog, F. Danielsson, A. Mardinoglu, A. Sivertsson, K. von Feilitzen, M. Forsberg, M. Zwahlen, I. Olsson, S. Navani, M. Huss, J. Nielsen, F. Ponten, M. Uhlén, Analysis of the human tissue-specific expression by genome-wide integration of transcriptomics and antibody-based proteomics. Mol Cell Proteomics 13, 397–406 (2014).

4. Y. Nakamura, T. Nguyen, N. Mor, C. J. Torio, H. Thulaseedharan, D. Dominissini, J. G. Gleeson, When loss is gain: truncating mutations in additional sex combs (ASXL) gene family in cancer and neurodevelopment. Trends Genet, S0168-9525(26)00038–7 (2026).

5. E. A. Medina, C. R. Delma, F.-C. Yang, ASXL1/2 mutations and myeloid malignancies. J Hematol Oncol 15, 127 (2022).

6. O. Abdel-Wahab, J. Gao, M. Adli, A. Dey, T. Trimarchi, Y. R. Chung, C. Kuscu, T. Hricik, D. Ndiaye-Lobry, L. M. Lafave, R. Koche, A. H. Shih, O. A. Guryanova, E. Kim, S. Li, S. Pandey, J. Y. Shin, L. Telis, J. Liu, P. K. Bhatt, S. Monette, X. Zhao, C. E. Mason, C. Y. Park, B. E. Bernstein, I. Aifantis, R. L. Levine, Deletion of Asxl1 results in myelodysplasia and severe developmental defects in vivo. J Exp Med 210, 2641–2659 (2013).

7. M. Balasubramanian, J. Willoughby, A. E. Fry, A. Weber, H. V. Firth, C. Deshpande, J. N. Berg, K. Chandler, K. A. Metcalfe, W. Lam, D. T. Pilz, S. Tomkins, Delineating the phenotypic spectrum of Bainbridge-Ropers syndrome: 12 new patients with de novo, heterozygous, loss-of-function mutations in ASXL3 and review of published literature. J Med Genet 54, 537–543 (2017).

8. A. Balasubramani, A. Larjo, J. A. Bassein, X. Chang, R. B. Hastie, S. M. Togher, H. Lähdesmäki, A. Rao, Cancer-associated ASXL1 mutations may act as gain-of-function mutations of the ASXL1-BAP1 complex. Nat Commun 6, 7307 (2015).

9. A. Manole, S. Efthymiou, E. O’Connor, M. I. Mendes, M. Jennings, R. Maroofian, I. Davagnanam, K. Mankad, M. R. Lopez, V. Salpietro, R. Harripaul, L. Badalato, J. Walia, C. S. Francklyn, A. Athanasiou-Fragkouli, R. Sullivan, S. Desai, K. Baranano, F. Zafar, N. Rana, M. Ilyas, A. Horga, M. Kara, F. Mattioli, A. Goldenberg, H. Griffin, A. Piton, L. B. Henderson, B. Kara, A. D. Aslanger, J. Raaphorst, R. Pfundt, R. Portier, M. Shinawi, A. Kirby, K. M. Christensen, L. Wang, R. O. Rosti, S. A. Paracha, M. T. Sarwar, D. Jenkins, SYNAPS Study Group, J. Ahmed, F. A. Santoni, E. Ranza, J. Iwaszkiewicz, C. Cytrynbaum, R. Weksberg, I. M. Wentzensen, M. J. Guillen Sacoto, Y. Si, A. Telegrafi, M. V. Andrews, D. Baldridge, H. Gabriel, J. Mohr, B. Oehl-Jaschkowitz, S. Debard, B. Senger, F. Fischer, C. van Ravenwaaij, A. J. M. Fock, S. J. C. Stevens, J. Bähler, A. Nasar, J. F. Mantovani, A. Manzur, A. Sarkozy, D. E. C. Smith, G. S. Salomons, Z. M. Ahmed, S. Riazuddin, S. Riazuddin, M. A. Usmani, A. Seibt, M. Ansar, S. E. Antonarakis, J. B. Vincent, M. Ayub, M. Grimmel, A. M. Jelsig, T. D. Hjortshøj, H. G. Karstensen, M. Hummel, T. B. Haack, Y. Jamshidi, F. Distelmaier, R. Horvath, J. G. Gleeson, H. Becker, J.-L. Mandel, D. A. Koolen, H. Houlden, De Novo and Bi-allelic Pathogenic Variants in NARS1 Cause Neurodevelopmental Delay Due to Toxic Gain-of-Function and Partial Loss-of-Function Effects. Am J Hum Genet 107, 311–324 (2020).

10. D. Nagy, S. Verheyen, K. M. Wigby, A. Borovikov, A. Sharkov, V. Slegesky, A. Larson, C. Fagerberg, C. Brasch-Andersen, M. Kibæk, I. Bader, R. Hernan, F. A. High, W. K. Chung, J. H. Schieving, J. Behunova, M. Smogavec, F. Laccone, M. Witsch-Baumgartner, J. Zobel, H.-C. Duba, D. Weis, Genotype-Phenotype Comparison in POGZ-Related Neurodevelopmental Disorders by Using Clinical Scoring. Genes (Basel) 13, 154 (2022).

11. C. C. Y. Mak, D. Doherty, A. E. Lin, N. Vegas, M. T. Cho, G. Viot, C. Dimartino, J. D. Weisfeld-Adams, D. Lessel, S. Joss, C. Li, C. Gonzaga-Jauregui, Y. A. Zarate, N. Ehmke, D. Horn, C. Troyer, S. G. Kant, Y. Lee, G. E. Ishak, G. Leung, A. Barone Pritchard, S. Yang, E. G. Bend, F. Filippini, C. Roadhouse, N. Lebrun, M. G. Mehaffey, P.-M. Martin, B. Apple, F. Millan, O. Puk, M. J. V. Hoffer, L. B. Henderson, R. McGowan, I. M. Wentzensen, S. Pei, F. R. Zahir, M. Yu, W. T. Gibson, A. Seman, M. Steeves, J. R. Murrell, S. Luettgen, E. Francisco, T. M. Strom, L. Amlie-Wolf, A. M. Kaindl, W. G. Wilson, S. Halbach, L. Basel-Salmon, N. Lev-El, J. Denecke, L. E. L. M. Vissers, K. Radtke, J. Chelly, E. Zackai, J. M. Friedman, M. J. Bamshad, D. A. Nickerson, University of Washington Center for Mendelian Genomics, R. R. Reid, K. Devriendt, J.-H. Chae, E. Stolerman, C. McDougall, Z. Powis, T. Bienvenu, T. Y. Tan, N. Orenstein, W. B. Dobyns, J. T. Shieh, M. Choi, D. Waggoner, K. W. Gripp, M. J. Parker, J. Stoler, S. Lyonnet, V. Cormier-Daire, D. Viskochil, T. L. Hoffman, J. Amiel, B. H. Y. Chung, C. T. Gordon, MN1 C-terminal truncation syndrome is a novel neurodevelopmental and craniofacial disorder with partial rhombencephalosynapsis. Brain 143, 55–68 (2020).

12. A. Behera, G. K. Panigrahi, A. Sahoo, Nonsense-Mediated mRNA Decay in Human Health and Diseases: Current Understanding, Regulatory Mechanisms and Future Perspectives. Mol Biotechnol 67, 3374–3390 (2025).

13. M. A. Cortázar, J. Schmidt, I. Egab, Z. Coban-Akdemir, S. Jagannathan, Genomic stop codon scanning reveals quantitative principles of nonsense-mediated mRNA decay. bioRxiv, 2025.12.20.695734 (2025).

14. H. Yang, S. Kurtenbach, Y. Guo, I. Lohse, M. A. Durante, J. Li, Z. Li, H. Al-Ali, L. Li, Z. Chen, M. G. Field, P. Zhang, S. Chen, S. Yamamoto, Z. Li, Y. Zhou, S. D. Nimer, J. W. Harbour, C. Wahlestedt, M. Xu, F.-C. Yang, Gain of function of ASXL1 truncating protein in the pathogenesis of myeloid malignancies. Blood 131, 328–341 (2018).

15. Z. Dong, H. Sepulveda, L. J. Arteaga-Vazquez, C. Blouin, J. Fernandez, M. Binder, W.-C. Chou, H.-F. Tien, M. M. Patnaik, G. J. Faulkner, S. A. Myers, A. Rao, A mutant ASXL1-BAP1-EHMT complex contributes to heterochromatin dysfunction in clonal hematopoiesis and chronic monomyelocytic leukemia. Proc Natl Acad Sci U S A 122, e2413302121 (2025).

19. H. P. Tirumala, L. Wang, Y. Li, S. S. Bajikar, A. G. Anderson, W. Wang, A. J. Trostle, M. Zahabiyon, A. Bajic, J. J. Kim, H. Chen, Z. Liu, H. Y. Zoghbi, Modulating alternative splicing of MECP2 is a potential therapeutic strategy for Rett syndrome. Sci Transl Med 18, eadq4529 (2026).

20. A. Campagne, M.-K. Lee, D. Zielinski, A. Michaud, S. Le Corre, F. Dingli, H. Chen, L. Z. Shahidian, I. Vassilev, N. Servant, D. Loew, E. Pasmant, S. Postel-Vinay, M. Wassef, R. Margueron, BAP1 complex promotes transcription by opposing PRC1-mediated H2A ubiquitylation. Nat Commun 10, 348 (2019).

21. J. Jin, X. L. Ang, T. Shirogane, J. Wade Harper, Identification of substrates for F-box proteins. Methods Enzymol 399, 287–309 (2005).

22. N. Tsuboyama, A. P. Szczepanski, Z. Zhao, L. Wang, MBD5 and MBD6 stabilize the BAP1 complex and promote BAP1-dependent cancer. Genome Biol 23, 206 (2022).

23. I. Koren, R. T. Timms, T. Kula, Q. Xu, M. Z. Li, S. J. Elledge, The Eukaryotic Proteome Is Shaped by E3 Ubiquitin Ligases Targeting C-Terminal Degrons. Cell 173, 1622–1635.e14 (2018).

24. Z. Lu, S. Xu, C. Joazeiro, M. H. Cobb, T. Hunter, The PHD domain of MEKK1 acts as an E3 ubiquitin ligase and mediates ubiquitination and degradation of ERK1/2. Mol Cell 9, 945–956 (2002).

25. C. Hou, Y. Li, M. Wang, H. Wu, T. Li, Systematic prediction of degrons and E3 ubiquitin ligase binding via deep learning. BMC Biol 20, 162 (2022).

26. n-Lorem Foundation, An Open-Label Single Center, Single Participant Study of an Experimental Antisense Oligonucleotide Treatment for Bainbridge-Ropers Syndrome Due to ASXL3 Gene Variant (clinicaltrials.gov, 2025; https://clinicaltrials.gov/study/NCT07197268).

27. N. Miyake, H. Takahashi, K. Nakamura, B. Isidor, Y. Hiraki, E. Koshimizu, M. Shiina, K. Sasaki, H. Suzuki, R. Abe, Y. Kimura, T. Akiyama, S.-I. Tomizawa, T. Hirose, K. Hamanaka, S. Miyatake, S. Mitsuhashi, T. Mizuguchi, A. Takata, K. Obo, M. Kato, K. Ogata, N. Matsumoto, Gain-of-Function MN1 Truncation Variants Cause a Recognizable Syndrome with Craniofacial and Brain Abnormalities. Am J Hum Genet 106, 13–25 (2020).

28. J. White, J. F. Mazzeu, A. Hoischen, S. N. Jhangiani, T. Gambin, M. C. Alcino, S. Penney, J. M. Saraiva, H. Hove, F. Skovby, H. Kayserili, E. Estrella, A. T. Vulto-van Silfhout, M. Steehouwer, D. M. Muzny, V. R. Sutton, R. A. Gibbs, Baylor-Hopkins Center for Mendelian Genomics, J. R. Lupski, H. G. Brunner, B. W. M. van Bon, C. M. B. Carvalho, DVL1 frameshift mutations clustering in the penultimate exon cause autosomal-dominant Robinow syndrome. Am J Hum Genet 96, 612–622 (2015).

29. S. Jansen, S. Geuer, R. Pfundt, R. Brough, P. Ghongane, J. C. Herkert, E. J. Marco, M. H. Willemsen, T. Kleefstra, M. Hannibal, J. T. Shieh, S. A. Lynch, F. Flinter, D. R. FitzPatrick, A. Gardham, B. Bernhard, N. Ragge, R. Newbury-Ecob, R. Bernier, M. Kvarnung, E. A. H. Magnusson, M. W. Wessels, M. A. van Slegtenhorst, K. G. Monaghan, P. de Vries, J. A. Veltman, Deciphering Developmental Disorders Study, C. J. Lord, L. E. L. M. Vissers, B. B. A. de Vries, De Novo Truncating Mutations in the Last and Penultimate Exons of PPM1D Cause an Intellectual Disability Syndrome. Am J Hum Genet 100, 650–658 (2017).

30. M. Veiner, I. Toledano, G. Palou-Márquez, B. Lehner, F. Supek, Quantitative prediction of nonsense-mediated mRNA decay across human genes by genomic language model and large-scale mutational scanning, 2026.03.24.714003 (2026).

31. X. Robin, N. Turck, A. Hainard, N. Tiberti, F. Lisacek, J.-C. Sanchez, M. Müller, pROC: an open-source package for R and S+ to analyze and compare ROC curves. BMC Bioinformatics 12, 77 (2011).

32. I. Tang, A. Nisal, A. Reed, T. B. Ware, A. Johansen, M. S. Zaki, B. F. Cravatt, J. G. Gleeson, Lipidomic profiling of mouse brain and human neuron cultures reveals a role for Mboat7 in mTOR-dependent neuronal migration. Sci Transl Med 17, eadp5247 (2025).

33. L. Wang, Y. Nakamura, J. Li, D. Sievert, Y. Liu, T. Nguyen, P. S. Jetti, E. Thai, R. Y. Zhou, J. Weng, N. Meave, M. Yadavilli, R. Howarth, K. Camey, N. Banka, C. Owusu-Hammond, C. Barrows, S. F. Kingsmore, M. S. Zaki, E. Mukamel, J. G. Gleeson, A phenotypic brain organoid atlas and biobank for neurodevelopmental disorders. Cell Stem Cell 32, 1923–1940.e7 (2025).

34. S. A. Bustin, V. Benes, J. A. Garson, J. Hellemans, J. Huggett, M. Kubista, R. Mueller, T. Nolan, M. W. Pfaffl, G. L. Shipley, J. Vandesompele, C. T. Wittwer, The MIQE guidelines: minimum information for publication of quantitative real-time PCR experiments. Clin Chem 55, 611–622 (2009).

35. B. A. Anderson, G. C. Freestone, A. Low, C. L. De-Hoyos, W. J. D. Iii, M. E. Østergaard, M. T. Migawa, M. Fazio, W. B. Wan, A. Berdeja, E. Scandalis, S. A. Burel, T. A. Vickers, S. T. Crooke, E. E. Swayze, X. Liang, P. P. Seth, Towards next generation antisense oligonucleotides: mesylphosphoramidate modification improves therapeutic index and duration of effect of gapmer antisense oligonucleotides. Nucleic Acids Res 49, 9026–9041 (2021).

36. A. Korff, X. Yang, O. Ozdemir, A. Samanta, Y.-D. Wang, T. Patni, A. J. Lavado, A. M. Kavirayani, J. Ochaba, B. Powers, C. F. Bennett, H. J. Kim, J. P. Taylor, Preclinical evaluation of antisense oligonucleotide therapy in a mouse model of HNRNPH2-related neurodevelopmental disorder. bioRxiv, 2025.11.04.686541 (2025).

37. T. F. Gendron, J. Chew, J. N. Stankowski, L. R. Hayes, Y.-J. Zhang, M. Prudencio, Y. Carlomagno, L. M. Daughrity, K. Jansen-West, E. A. Perkerson, A. O’Raw, C. Cook, L. Pregent, V. Belzil, M. van Blitterswijk, L. J. Tabassian, C. W. Lee, M. Yue, J. Tong, Y. Song, M. Castanedes-Casey, L. Rousseau, V. Phillips, D. W. Dickson, R. Rademakers, J. D. Fryer, B. K. Rush, O. Pedraza, A. M. Caputo, P. Desaro, C. Palmucci, A. Robertson, M. G. Heckman, N. N. Diehl, E. Wiggs, M. Tierney, L. Braun, J. Farren, D. Lacomis, S. Ladha, C. N. Fournier, L. F. McCluskey, L. B. Elman, J. B. Toledo, J. D. McBride, C. Tiloca, C. Morelli, B. Poletti, F. Solca, A. Prelle, J. Wuu, J. Jockel-Balsarotti, F. Rigo, C. Ambrose, A. Datta, W. Yang, D. Raitcheva, G. Antognetti, A. McCampbell, J. C. Van Swieten, B. L. Miller, A. L. Boxer, R. H. Brown, R. Bowser, T. M. Miller, J. Q. Trojanowski, M. Grossman, J. D. Berry, W. T. Hu, A. Ratti, B. J. Traynor, M. D. Disney, M. Benatar, V. Silani, J. D. Glass, M. K. Floeter, J. D. Rothstein, K. B. Boylan, L. Petrucelli, Poly(GP) proteins are a useful pharmacodynamic marker for C9ORF72-associated amyotrophic lateral sclerosis. Sci Transl Med 9, eaai7866 (2017).

